# Clinical characteristics of COVID-19 in children and adolescents: a systematic review and meta-analysis

**DOI:** 10.1101/2021.03.12.21253472

**Authors:** Lixiang Lou, Hui Zhang, Baoming Tang, Ming Li, Zeqing Li, Haifang Cao, Jian Li, Yuliang Chong, Zhaowei Li

**Author notes:** These authors contributed equally to the manuscript (Lixiang Lou, Hui Zhang). **Corresponding Author**: Zhaowei Li, Dr Department of Traumatology orthopedics, Affiliated Hospital of Qinghai University, Xining, China, **Address**: 29 Tongren Road, Xining, Qinghai, China, **Postal code**:810000, Zhaowei Li. **Competing interests**: No authors have competing interests in this research. **Funding**: This research did not receive any specific grant from funding agencies in the public, commercial, or not-for-profit sectors. Contributors: All the authors designed the study. LL, HZ, ZL and BT designed the literature search and searched the articles. LL, HZ, ML, YC, JL, and BT contributed to statistical analysis, interpretation of data and data review. LL, HZ wrote the first draft of the article. All the authors revised the article and approved the final version.

## Abstract

**Background:** Although the number of COVID-19 (coronavirus disease 2019) cases continues to increase globally, there are few studies on the clinical characteristics of children and adolescents with COVID-19.

**Objective:** To conduct a comprehensive systematic evaluation and meta-analysis of the clinical characteristics of COVID-19 in children and adolescents to better guide the response to the current epidemic.

**Methods:** We searched PubMed, Embase, the Cochrane Library, Web of Science, CNKI (Chinese database), Clinical Trials.gov and chictr.org.cn (China). The methodological quality of the included literature was evaluated using the Quality Assessment Tool for Case Series Studies. Meta-analysis was performed using STATA 14.0. Heterogeneity was assessed by the Q statistic and quantified using I^2^. We used fixed-effects or random-effects models to pool clinical data in the meta-analysis. Publication bias was evaluated by the Begg’s test.

**Results:** We analyzed 49 studies involving 1627 patients. In the pooled data, the most common clinical symptoms were fever (56% [0.50−0.61]) and cough (45% [0.39−0.51]). The most common laboratory abnormalities were elevated procalcitonin (40% [0.23−0.57]), elevated lactate dehydrogenase (31% [0.19−0.43]), increased lymphocyte count (28% [0.17−0.42]), increased creatine kinase (28% [0.18− 0.40]), and elevated C-reactive protein (26% [0.17−0.36]). The most common abnormalities determined by computed tomography were lower-lobe involvement (56% [0.42-0.70]), ground-glass opacities (33% [0.25−0.42]), bilateral pneumonia (32% [0.24-0.40]), patchy shadowing (31% [0.18-0.45]), and upper lobe involvement (30% [0.20-0.41]).

**Conclusion:** Disease severity among children and adolescents with COVID-19 was milder than that among adult patients, with a greater proportion of mild and asymptomatic cases, and thus, the diagnosis of COVID-19 and control of the infection source are more challenging.

## Introduction

Severe acute respiratory syndrome coronavirus 2 (SARS-CoV-2) is a newly discovered coronavirus responsible for the coronavirus disease 2019 (COVID-19), which was first reported in Wuhan City, Hubei Province in December 2019 [1]. On January 12, 2020, the World Health Organization (WHO) officially named the novel coronavirus “2019 novel coronavirus (2019-nCoV)” [2]. So far, over 20□million people live with COVID-19 worldwide, with 750,000 deaths, and the death toll is still rising. Currently, COVID-19 outbreaks are continuing to spread across the globe. SARS-CoV-2 is spread by between-human transmission via droplets or close contact [3], but there are also studies showing that it can be transmitted from person to person by aerosols [4]. Fecal-oral routes may also constitute a potential person-to-person transmission route. Several studies have reported that the virus may still be transmitted from COVID-19-positive stool samples even after COVID-19 with negative detection of viral ribonucleic acid from nasopharyngeal swabs [5,13]. The development of the immune system in children and adolescents is not yet perfect, and it is easy for various types of viruses to cause infections. COVID-19 is a novel infectious disease. The population lacks immunity and is generally susceptible in crowded conditions. However, a study has found that COVID-19 in children below the age of 10 years constituted only 0.35% of cases [6].It is essential to identify and control patients who are suspected of having COVID-19 as early as possible and to prevent the further spread of the epidemic by controlling the source of infection and cutting off transmission pathways. Although the number of COVID-19 cases continues to grow globally, there are few studies focusing on the clinical manifestations, laboratory tests, chest imaging examinations, and complications for children and adolescents with COVID-19. We conducted a systematic review and meta-analysis of the clinical characteristics of children and adolescents to describe comprehensive characteristics of COVID-19 in children and adolescents to better treat and control the current outbreak.

## 1 Methods

### 1.1 Retrieval of published studies

PRISMA and MOOSE guidelines were used to conduct this systematic review and meta-analysis [7–8]. We used the Medical Subject Heading (MeSH) terms and corresponding keywords to develop the search strategy. The search terms included: “Child (MeSH)” or “children” or “Adolescent (MeSH)” or “Teens” and “COVID-19 (MeSH)” or “SARS-CoV-2 infection” and “clinical characteristics” or “clinical features”. We searched PubMed, Embase, the Cochrane Library, Web of Science, and CNKI (Chinese database). We also searched Clinical Trials.gov and chictr.org.cn (China) to find unpublished studies. There were no restrictions on use, including no language restrictions. The search time limit was from the establishment of each database to July 10, 2020. To ensure comprehensiveness of the search strategy, the search tasks were performed independently by two researchers (LL, BT). Details of the search strategy are provided as shown in Supplementary Text S1.

### 1.2 Inclusion and exclusion criteria

#### 1.2.1 Inclusion criteria

□ Research type: Not limited by research type or research design. □ Research object: Participants included children and adolescents between the ages of 1 and 18 years, cases with positive nucleic acid tests in patients with COVID-19. □Intervention was unrestricted. □ Outcomes included clinical manifestations, laboratory examinations, and CT examinations.

#### 1.2.2 Exclusion criteria

□ Infants less than 1 year of age. □ Fewer than 4 cases reported. □ Surveillance reports, community outbreaks, review papers, commentary articles, conference papers, duplicated reports, reports for which the full text cannot be obtained. □ There is no definite measure or laboratory marker for the diagnosis of COVID-19. □ The outcome was not clear.

### 1.3 Data extraction

Two researchers (HZ, LL) independently screened and cross-checked the literature regarding the inclusion and exclusion criteria. Any disagreement was resolved by discussion between the two reviewers. A customized form was used to record the first author, publication time, design of the trial, country, the number of COVID-19 patients, ages, sexes, durations between onset of symptoms and hospitalization (days), times of patient examinations, diagnostic criteria for COVID-19, clinical symptoms, laboratory outcomes, chest CT findings, and supportive treatment. In addition, we screened them according to whether blood or respiratory samples were positive by real-time polymerase chain reaction tests for SARS-CoV-2 nucleic acid.

### 1.4 Quality assessment of included meta-analyses

For the case series study, we used the Quality Assessment Tool for Case Series Studies to evaluate article suitability for systematic review [9]. Each item was scored as either 0 or 1 point based on the scoring criteria, and scores were summed across items to generate an overall quality score ranging from 0 to 9. Overall scores were classified as low risk (≥7 points), moderate risk (5–6 points), or high risk (0–4 points). Two evaluators (HZ, LL) separately performed quality assessment. Any disagreements were resolved through discussion among the investigators.

### 1.5 Statistical analysis

Meta-analysis was performed using STATA 14.0. A random-effects model was used to compute the combined prevalence and 95% confidence intervals (CI). We calculated the pooled incidence of clinical symptoms, laboratory findings, and chest CT findings. Heterogeneity was assessed using Cochran’s Q test with a significance level of p < 0.1 and quantified using the I^2^ statistic. Heterogeneity was quantified using the I-squared (*I*^*2*^) statistic and categorized as low (*I*^*2*^ <25%), moderate (*I*^*2*^=25-50%) or high (*I*^*2*^ >50%). A model of fixed effect (FE) was set with 95% CI if no statistical evidence of heterogeneity existed (*p*≥ 0.1, *I*^*2*^ ≤ 50%), while a model of random effect (RE) was applied to estimate pooled effect with 95% confidence intervals (CI) if statistical heterogeneity was found (*p* < 0.1, *I*^*2*^ > 50%). If significant heterogeneity (*p*< 0.1, *I*^*2*^ > 75%) was detected, we used sensitivity analysis to explore its possible sources. A random-effects model was used if the reason for heterogeneity could not be identified. Publication bias was detected using Begg’s test. Probability (*p*) values less than 0.05 were considered statistically significant.

## 2 Results

### 2.1 Systematic search results

The search strategy initially identified 467 publications, including 60 from PubMed, 131 from Embase, 5 from Cochrane, and 196 from Web of Science. Search results from different sources were entered into an EndNote library, from which 146 duplicates and 366 irrelevant studies were removed. Among the remaining 101 articles selected for full-text reading, 52 studies were removed. Finally, 49 studies [10–58] were included in the meta-analysis involving 1627 participants. Only three of 49 included studies reported critically ill patients [15,26,31]. These critically ill patients were not excluded from the meta-analysis. Mechanical ventilation analysis included only critically ill patients. We extracted data based on different outcome indicators and then conducted a single-arm meta-analysis. Figure 1 illustrates the detailed process of the literature review.

**Figure 1.**
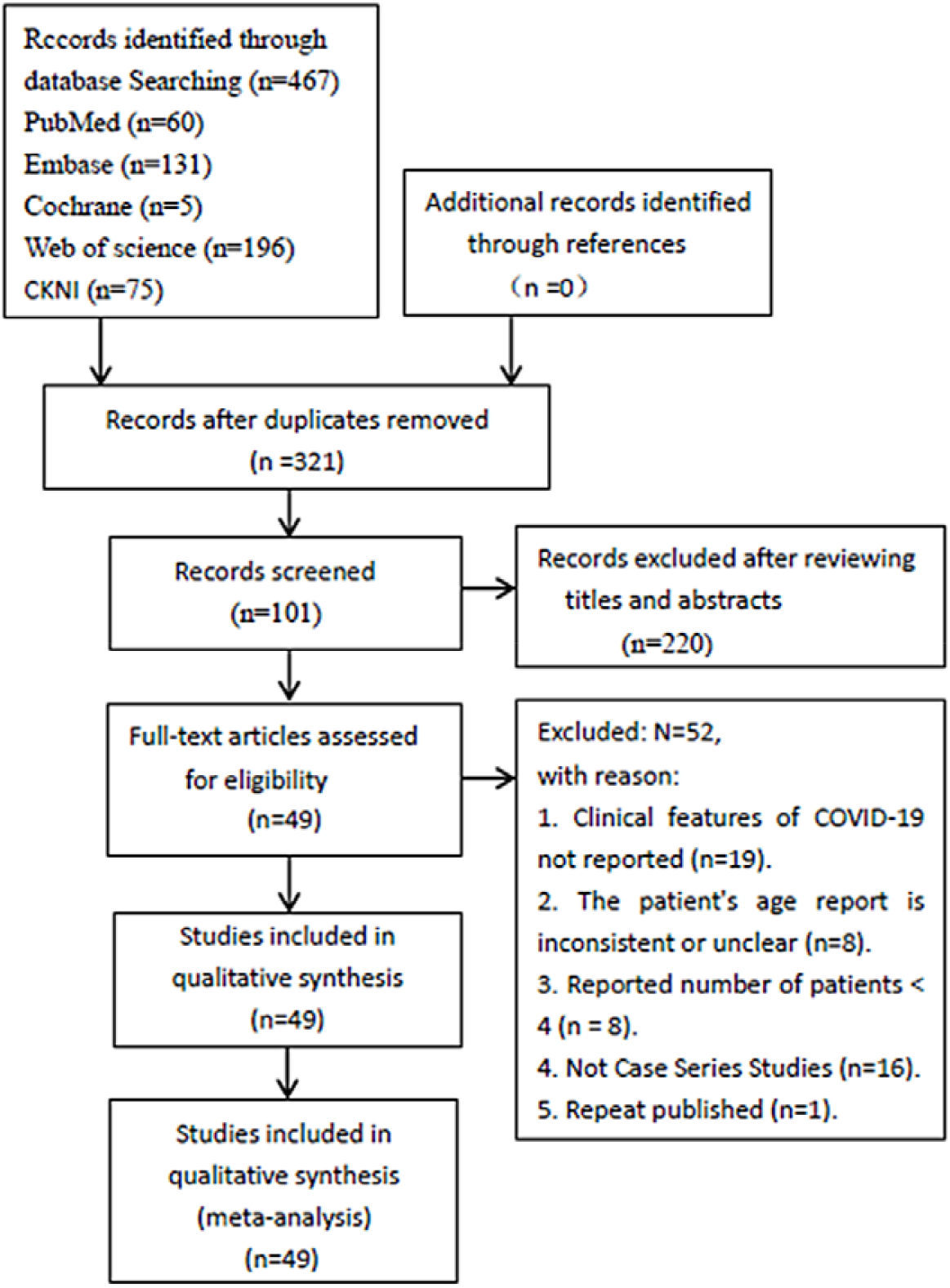
Flow chart of literature search and selection of studies.

### 2.2 Study characteristics

The characteristics of the studies are displayed in Table 1. The studies included were published between February 26, 2020 and June 29, 2020. Fifty articles including 41 (84%) English papers and 8 (16%) Chinese papers were included in our study, and all were journal articles. Patients were from six countries: 41 from China, 4 from the United States, 1 from Italy, 1 from France, 1 from Turkey, and 1 from Iran. All included studies were retrospective studies; one was a case-control study, and the remainder were retrospective case series. The number of patients enrolled in each study ranged from 4 to 157, and patients’ ages ranged from 1 to 18 years. Male patients accounted for 53%, and female patients accounted for 47%. All patients were confirmed to be positive for SARS-CoV-2 nucleic acid by respiratory or blood sample.

**Table 1.**
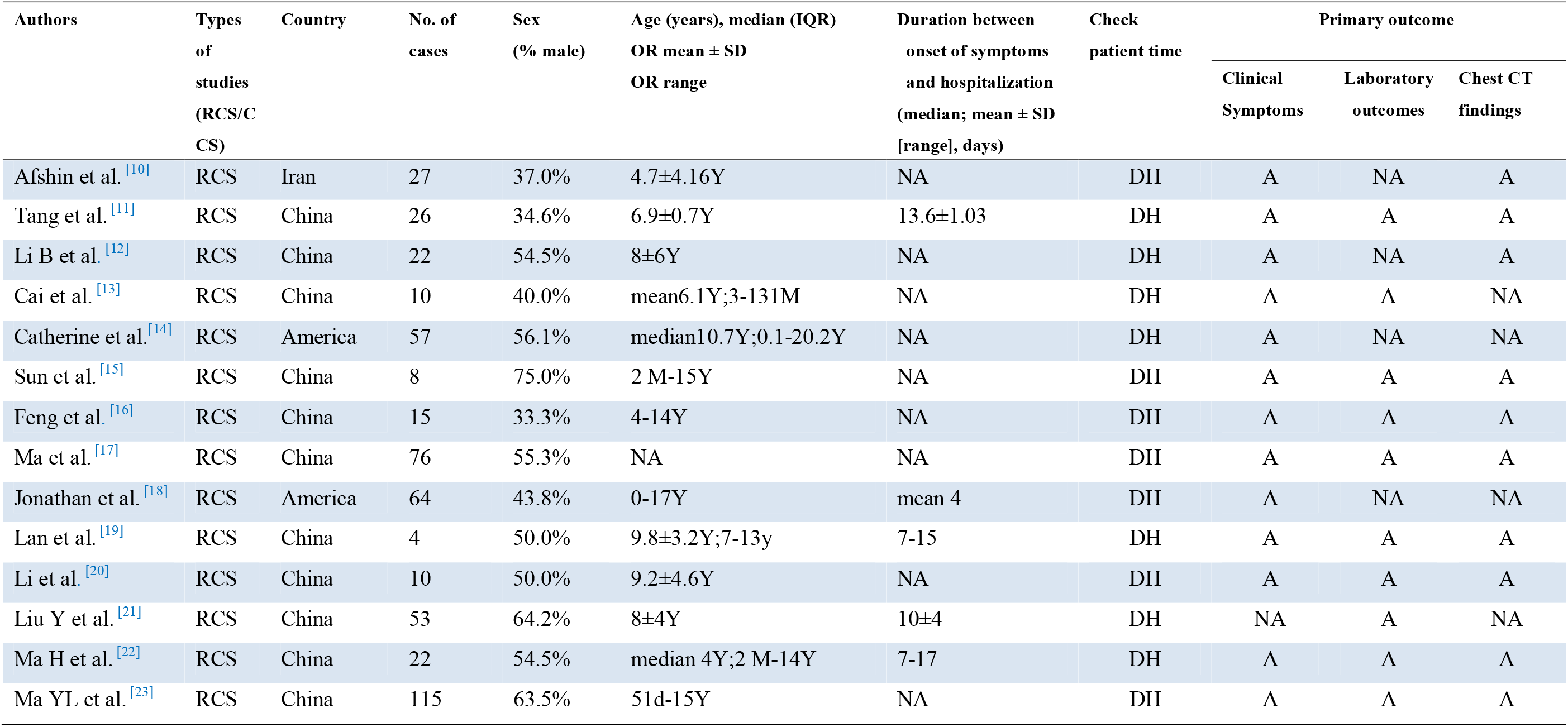
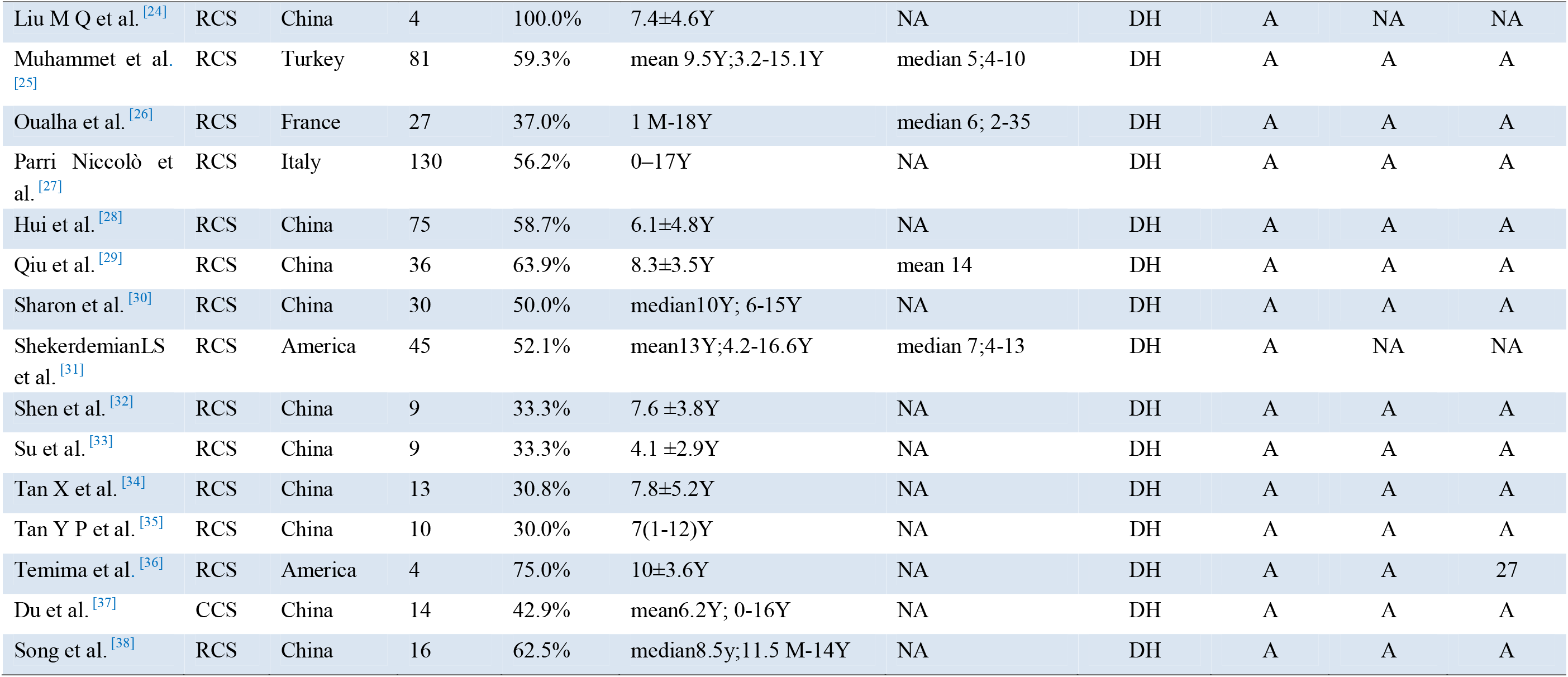
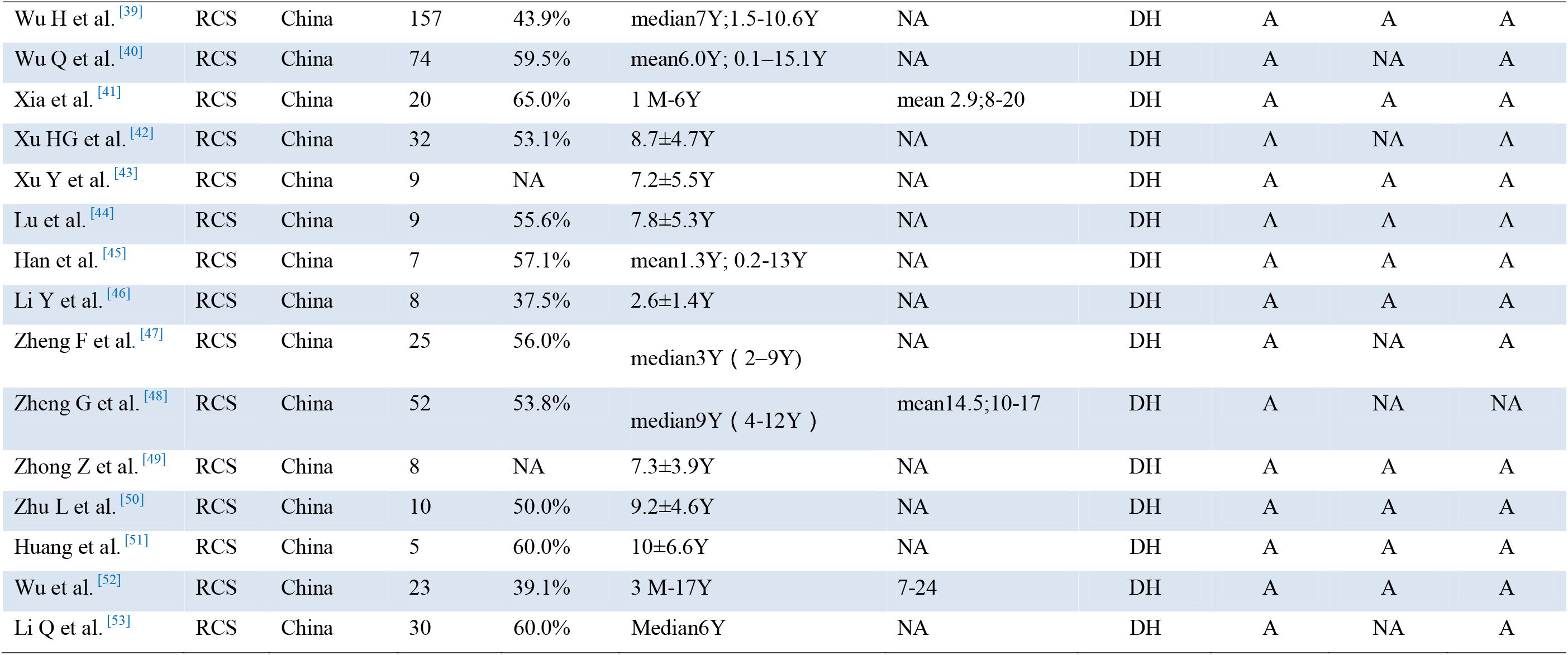
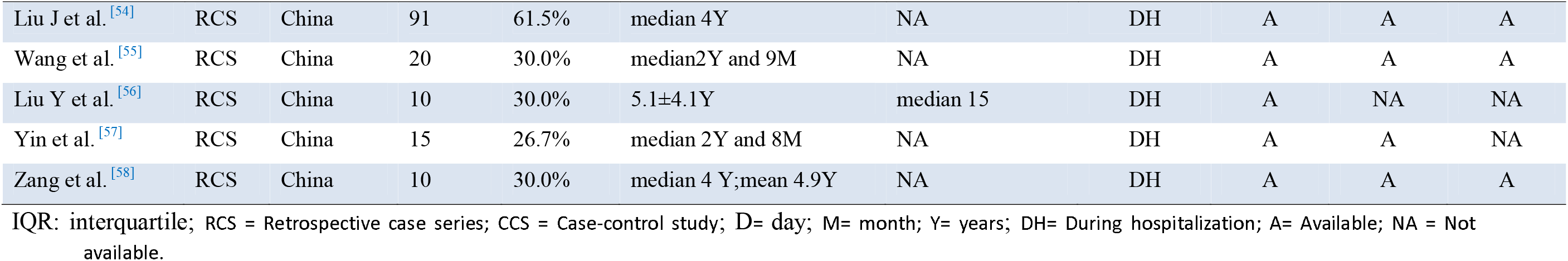
Characteristics of studies included in the meta-analysis.

### 2.3 Methodological assessment of the included literature

For the evidence of effectiveness meta-analysis, we used the Quality Assessment Tool for Case Series Studies to evaluate article suitability for the systematic review. Among the 49 studies included, 43 studies were rated as low risk of bias, 6 studies as moderate, and none were rated as high risk of bias (Table 2).

**Table 2.** Quality Assessment Tool for Case Series Studies.

| Author | NIH Quality Assessment Tool for Case Series Studies |  |  |  |  |  |  |  |  | Quality (Total Quality Score) |
| --- | --- | --- | --- | --- | --- | --- | --- | --- | --- | --- |
|  | Q1 | Q2 | Q3 | Q4 | Q5 | Q6 | Q7 | Q8 | Q9 |  |
| Afshin et al. <sup>[10]</sup> | 1 | 1 | 1 | NA | 1 | 1 | 1 | 1 | 1 | Good (8) |
| Tang et al. <sup>[11]</sup> | 1 | 1 | 1 | NA | 1 | 0 | 1 | 1 | 1 | Good (7) |
| Li B et al. <sup>[12]</sup> | 1 | 1 | 1 | NA | 1 | 1 | 1 | 1 | 1 | Good (8) |
| Cai et al. <sup>[13]</sup> | 1 | 1 | 1 | NA | 1 | 0 | 1 | 1 | 1 | Good (7) |
| Catherine et al. <sup>[14]</sup> | 1 | 0 | 1 | NA | 1 | 1 | 1 | 1 | 1 | Good (7) |
| Sun et al. <sup>[15]</sup> | 1 | 1 | 1 | NA | 1 | 1 | 1 | 1 | 1 | Good (8) |
|  | Q1 | Q2 | Q3 | Q4 | Q5 | Q6 | Q7 | Q8 | Q9 |  |
| Feng et al. <sup>[16]</sup> | 1 | 0 | 1 | NA | NR | 0 | 1 | 1 | 1 | Fair (5) |
| Ma et al. <sup>[17]</sup> | 1 | 0 | 1 | NA | NR | 0 | 1 | 1 | 1 | Fair (5) |
| Jonathan et al. <sup>[18]</sup> | 1 | 1 | 1 | NA | NR | 1 | 1 | 1 | 1 | Good (7) |
| Lan et al. <sup>[19]</sup> | 1 | 1 | 1 | NA | 1 | 1 | 1 | 1 | 1 | Good (8) |
| Li et al. <sup>[20]</sup> | 1 | 1 | 1 | NA | 1 | 1 | 1 | 1 | 1 | Good (8) |
| Liu Y et al. <sup>[21]</sup> | 1 | 1 | 1 | NA | 1 | 1 | 1 | 1 | 1 | Good (8) |
| Ma H et al. <sup>[22]</sup> | 1 | 1 | 1 | NA | 0 | 1 | 1 | 1 | 1 | Good (7) |
| Ma Y L et al. <sup>[23]</sup> | 1 | 1 | 0 | NA | NR | 0 | 1 | 1 | 1 | Fair (5) |
| Liu M Q et al. <sup>[24]</sup> | 1 | 1 | NR | NA | NR | 1 | 1 | 1 | 1 | Fair (6) |
| Muhammet et al. <sup>[25]</sup> | 1 | 1 | 1 | NA | 1 | 1 | 1 | 1 | 1 | Good (8) |
| Oualha et al. <sup>[26]</sup> | 1 | 1 | 1 | NA | 1 | 0 | 1 | 1 | 1 | Good (7) |
| Parri Niccolò et al. <sup>[27]</sup> | 1 | 1 | 1 | NA | 1 | 0 | 1 | 1 | 1 | Good (7) |
| Hui et al. <sup>[28]</sup> | 1 | 1 | 1 | NA | 1 | 1 | 1 | 1 | 1 | Good (8) |
| Qiu et al. <sup>[29]</sup> | 1 | 1 | 1 | NA | NR | 1 | 1 | 1 | 1 | Good (7) |
| Sharon et al. <sup>[30]</sup> | 1 | 1 | 1 | NA | NR | 1 | 1 | 1 | 1 | Good (7) |
| ShekerdemianLS et al. <sup>[31]</sup> | 1 | 1 | 1 | NA | 1 | 1 | 1 | 1 | 1 | Good (8) |
| Shen et al. <sup>[32]</sup> | 1 | 1 | 1 | NA | 1 | 1 | 1 | 1 | 1 | Good (8) |
|  | Q1 | Q2 | Q3 | Q4 | Q5 | Q6 | Q7 | Q8 | Q9 |  |
| Su et al. <sup>[33]</sup> | 1 | 1 | 1 | NA | NR | 1 | 1 | 1 | 1 | Good (7) |
| Tan X et al. <sup>[34]</sup> | 1 | 1 | 1 | NA | 1 | 1 | 1 | 1 | 1 | Good (8) |
| Tan Y P et al. <sup>[35]</sup> | 1 | 1 | 1 | NA | 1 | 1 | 1 | 1 | 1 | Good (8) |
| Temima et al. <sup>[36]</sup> | 1 | 1 | 1 | NA | 1 | 1 | 1 | 1 | 1 | Good (8) |
| Du et al. <sup>[37]</sup> | 1 | 1 | 1 | 1 | NR | 1 | 1 | 1 | 1 | Good (8) |
| Song et al. <sup>[38]</sup> | 1 | 1 | 1 | NA | 1 | 1 | 1 | 1 | 1 | Good (8) |
| Wu H et al. <sup>[39]</sup> | 1 | 1 | 1 | NA | NR | 0 | 1 | 1 | 1 | Fair (6) |
| Wu Q et al. <sup>[40]</sup> | 1 | 1 | 1 | NA | 1 | 1 | 1 | 1 | 1 | Good (8) |
| Xia et al. <sup>[41]</sup> | 1 | 0 | 1 | NA | 0 | 1 | 1 | 1 | 1 | Fair (6) |
| Xu HG et al. <sup>[42]</sup> | 1 | 1 | 1 | NA | 1 | 1 | 1 | 1 | 1 | Good (8) |
| Xu Y et al. <sup>[43]</sup> | 1 | 1 | 1 | NA | NR | 1 | 1 | 1 | 1 | Good (7) |
| Lu et al. <sup>[44]</sup> | 1 | 1 | 1 | NA | NR | 1 | 1 | 1 | 1 | Good (7) |
| Han et al. <sup>[45]</sup> | 1 | 1 | 1 | NA | 1 | 1 | 1 | 1 | 1 | Good (8) |
| Li Y et al. <sup>[46]</sup> | 1 | 1 | 1 | NA | 1 | 1 | 1 | 1 | 1 | Good (8) |
| Zheng F et al. <sup>[47]</sup> | 1 | 0 | 1 | NA | 1 | 1 | 1 | 1 | 1 | Good (7) |
| Zheng G et al. <sup>[48]</sup> | 1 | 0 | 1 | NA | 1 | 1 | 1 | 1 | 1 | Good (7) |
| Zhong Z et al. <sup>[49]</sup> | 1 | 1 | 1 | NA | NR | 1 | 1 | 1 | 1 | Good (7) |
| Zhu L et al. <sup>[50]</sup> | 1 | 1 | 1 | NA | 1 | 1 | 1 | 1 | 1 | Good (8) |
|  | Q1 | Q2 | Q3 | Q4 | Q5 | Q6 | Q7 | Q8 | Q9 |  |
| Huang et al. <sup>[51]</sup> | 1 | 1 | 1 | NA | NR | 1 | 1 | 1 | 1 | Good (7) |
| Wu et al. <sup>[52]</sup> | 1 | 1 | 1 | NA | 1 | 0 | 1 | 1 | 1 | Good (7) |
| Li Q et al. <sup>[53]</sup> | 1 | 0 | 1 | NA | 1 | 1 | 1 | 1 | 1 | Good (7) |
| Liu J et al. <sup>[54]</sup> | 1 | 1 | 1 | NA | 1 | 1 | 1 | 1 | 1 | Good (8) |
| Wang et al. <sup>[55]</sup> | 1 | 0 | 1 | NA | 1 | 1 | 1 | 1 | 1 | Good (7) |
| Liu Y et al. <sup>[56]</sup> | 1 | 1 | 1 | NA | 1 | 1 | 1 | 1 | 1 | Good (8) |
| Yin et al. <sup>[57]</sup> | 1 | 1 | 1 | NA | 1 | 1 | 1 | 1 | 1 | Good (8) |
| Zang et al. <sup>[58]</sup> | 1 | 1 | 1 | NA | 1 | 1 | 1 | 1 | 1 | Good (8) |

Good: Met 7–9 criteria, Fair: Met 4–6 criteria, Poor: Met 0–3 criteria. NA = not applicable, NIH = National Institutes of Health, NR = not reported, Q1: Was study question or objective clearly stated?, Q2: Was study population clearly and fully described, including case definition?, Q3: Were cases consecutive?, Q4: Were subjects comparable?, Q5: Was intervention clearly described?, Q6: Were outcome measures clearly defined, valid, reliable, and implemented consistently across all study participants?, Q7: Was length of follow-up adequate?, Q8: Were statistical methods well-described?, Q9: Were results well-described?.

### 2.4 Meta-analysis results

We meta-analyzed the prevalence of 18 clinically relevant symptoms (Table 3), the prevalence of 12 laboratory outcomes (Table 4), and the prevalence of 9 chest CT findings (Table 5). The analysis of the 48 articles included showed that boys (53% [0.50−0.56]) had a higher prevalence of COVID-19 than did girls (47% [0.44−0.50]).

**Table 3.** Clinical Symptoms.

| Outcomes | No. includ-ed studies | No. events/total | I<sup>2</sup> | Q test P= | Fixed pooled | Random pooled | 95% Conf. Interval | Test of Z= | Test of P= | Begg's Test P= |
| --- | --- | --- | --- | --- | --- | --- | --- | --- | --- | --- |
| With clinical symptoms | 35 | 796/984 | 90.67% | 0.00 | NA | 0.86 | 0.77–0.93 | 18.71 | 0.00 | 0.158 |
| Fever | 44 | 729/1404 | 72.68% | 0.00 | NA | 0.56 | 0.50–0.61 | 25.06 | 0.00 | 0.418 |
| Cough | 43 | 664/1393 | 71.36% | 0.00 | NA | 0.45 | 0.39–0.51 | 21.90 | 0.00 | 0.509 |
| Sneezing | 13 | 111/627 | 80.58% | 0.00 | NA | 0.14 | 0.08–0.22 | 6.13 | 0.00 | 0.669 |
| Sore throat | 19 | 85/759 | 76.67% | 0.00 | NA | 0.10 | 0.05–0.16 | 5.84 | 0.00 | 0.039 |
| Increased sputum volume | 5 | 12/146 | 0.00% | 0.58 | 0.07 | NA | 0.03–0.13 | 4.81 | 0.00 | 0.806 |
| Anhelation | 14 | 70/607 | 84.96% | 0.00 | NA | 0.11 | 0.04–0.19 | 4.55 | 0.00 | 0.037 |
| Myalgia | 8 | 38/347 | 76.06% | 0.00 | NA | 0.08 | 0.02–0.17 | 3.36 | 0.00 | 0.386 |
| Fatigue | 15 | 99/755 | 75.04% | 0.00 | NA | 0.09 | 0.05–0.15 | 5.75 | 0.00 | 0.586 |
| Headache | 14 | 71/654 | 80.61% | 0.00 | NA | 0.10 | 0.04–0.17 | 4.79 | 0.00 | 0.042 |
| Abdominal Pain | 7 | 17/244 | 53.45% | 0.04 | NA | 0.06 | 0.01–0.12 | 3.08 | 0.00 | 0.133 |
| Diarrhea | 24 | 142/1100 | 87.06% | 0.00 | NA | 0.12 | 0.06–0.19 | 5.91 | 0.00 | 0.001 |
| Vomiting | 18 | 120/693 | 88.08% | 0.00 | NA | 0.14 | 0.06–0.24 | 4.86 | 0.00 | 0.010 |
| Renal insufficiency | 3 | 6/178 | 71.29% | 0.03 | NA | 0.03 | 0.00–0.11 | 1.89 | 0.06 | 0.296 |
| Mechanical ventilation | 3 | 30/78 | 0.00% | 0.96 | 0.38 | NA | 0.27–0.50 | 9.62 | 0.00 | 0.296 |
| Death | 49 | 7/1627 | 0.00% | 0.98 | 0.00 | NA | 0.00–0.00 | 0.00 | 1.00 | 0.000 |
| Percentage of boys | 47 | 852/1613 | 31.37% | 0.02 | 0.53 | NA | 0.50–0.56 | 41.17 | 0.00 | 0.229 |
| Proportion of girls | 47 | 761/1613 | 31.37% | 0.02 | 0.47 | NA | 0.44–0.50 | 38.86 | 0.00 | 0.229 |

**Table 4.** Laboratory outcomes.

| Outcomes | No.includ-ed studies | No. events/total | I<sup>2</sup> | Q test P= | Fixed pooled | Random pooled | 95% Conf. Interval | Test of Z= | Test of P= | Begg's Test P= |
| --- | --- | --- | --- | --- | --- | --- | --- | --- | --- | --- |
| Leukocytes ↑ | 14 | 37/335 | 70.45% | 0.00 | NA | 0.15 | 0.07–0.25 | 4.92 | 0.00 | 0.024 |

| Outcomes | No.includ-<br>ed studies | No.<br>events/total | I<sup>2</sup> | Q test<br>P= | Fixed<br>pooled | Random<br>pooled | 95% Conf.<br>Interval | Test of<br>Z= | Test of<br>P= | Begg's Test<br>P= |
| --- | --- | --- | --- | --- | --- | --- | --- | --- | --- | --- |
| Leukocytes ↓ | 24 | 135/645 | 78.23% | 0.00 | NA | 0.21 | 0.13–0.29 | 8.02 | 0.00 | 0.196 |
| lymphocyte ↑ | 19 | 126/454 | 84.64% | 0.00 | NA | 0.28 | 0.17–0.42 | 6.67 | 0.00 | 0.276 |
| lymphocyte ↓ | 19 | 88/554 | 66.42% | 0.00 | NA | 0.15 | 0.09–0.21 | 7.47 | 0.00 | 0.674 |
| ALT ↑ | 13 | 45/448 | 39.76% | 0.07 | 0.07 | NA | 0.05–0.10 | 6.11 | 0.00 | 0.032 |
| AST ↑ | 7 | 47/242 | 78.83% | 0.00 | NA | 0.24 | 0.09–0.43 | 4.27 | 0.00 | 0.095 |
| Creatinine ↑ | 2 | 4/121 | NA | NA | NA | NA | NA | NA | NA | NA |
| Creatine Kinase ↑ | 13 | 108/444 | 77.70% | 0.00 | NA | 0.28 | 0.18–0.40 | 7.88 | 0.00 | 0.271 |
| Lactate<br>dehydrogenase ↑ | 13 | 121/468 | 80.93% | 0.00 | NA | 0.31 | 0.19–0.43 | 7.47 | 0.00 | 0.299 |
| C reactive protein ↑ | 21 | 196/710 | 84.45% | 0.00 | NA | 0.26 | 0.17–0.36 | 8.33 | 0.00 | 0.880 |
| D - dimer ↑ | 9 | 35/234 | 68.53% | 0.00 | NA | 0.19 | 0.09–0.31 | 5.06 | 0.00 | 0.022 |
| Procalcitonin ↑ | 13 | 195/495 | 91.14% | 0.00 | NA | 0.40 | 0.23–0.57 | 6.58 | 0.00 | 0.951 |

ALT: Alanine aminotransferase; AST: Aspartate aminotransferase.

**Table 5.** Chest CT findings.

| Outcomes | No.<br>includ-ed<br>studies | No.<br>events/total | I<sup>2</sup> | Q test<br>P= | Fixed<br>pooled | Random<br>pooled | 95% Conf.<br>Interval | Test of<br>Z= | Test of<br>P= | Begg's Test<br>P= |
| --- | --- | --- | --- | --- | --- | --- | --- | --- | --- | --- |
| CT showed infection | 25 | 386/574 | 78.19% | 0.00 | NA | 0.67 | 0.58–0.76 | 17.32 | 0.00 | 0.453 |
| Bilateral pneumonia | 20 | 191/694 | 74.19% | 0.00 | NA | 0.32 | 0.24–0.40 | 11.80 | 0.00 | 0.019 |
| Unilateral pneumonia | 16 | 151/552 | 32.92% | 0.10 | 0.26 | NA | 0.22–0.30 | 14.62 | 0.00 | 0.892 |
| Involvement in upper lobe | 9 | 81/237 | 60.59% | 0.01 | NA | 0.30 | 0.20–0.41 | 8.79 | 0.00 | 0.208 |
| Involvement in lower lobe | 10 | 150/262 | 77.52% | 0.00 | NA | 0.56 | 0.42–0.70 | 10.56 | 0.00 | 0.857 |
| Patchy shadowing | 5 | 69/195 | 67.69% | 0.01 | NA | 0.31 | 0.18–0.45 | 6.92 | 0.00 | 0.221 |
| Ground-glass opacities | 27 | 311/970 | 84.44% | 0.00 | NA | 0.33 | 0.25–0.42 | 11.50 | 0.00 | 0.217 |
| Pleural fluid | 3 | 6/218 | 56.13% | 0.10 | NA | 0.03 | 0.00–0.08 | 2.18 | 0.03 | 0.296 |
| Tubercles | 4 | 13/93 | 33.52% | 0.21 | 0.13 | NA | 0.06–0.21 | 4.64 | 0.00 | 0.089 |

CT: Computed Tomography.

Among the 18 frequently reported clinical symptoms (Table 3), the incidence of clinical symptoms of COVID-19 in children and adolescents was 86% [95% CI 0.77−0.93], and the most common clinical symptoms were fever (56% [0.50−0.61]) (Supplementary Figs S 1), cough (45% [0.39−0.51]) (Supplementary Figs S 2), sneezing (14% [0.08−0.22]), and vomiting (14% [0.06−0.24]). Other symptoms included diarrhea (12% [0.06−0.19]), anhelation (11% [0.04−0.19]), sore throat (10% [0.05−0.16]), headache (10% [0.04−0.17]), fatigue (9% [0.05 −0.15]), myalgia (8% [0.02−0.17]), increased sputum volume (7% [0.03−0.13]), and abdominal pain (6% [0.01−0.12]). In addition, the meta-analysis included 3 studies with critically ill children and showed that the incidence of mechanical ventilation in severely ill children was 38% [0.27−0.50]. All the above results were statistically significant (p□<□0.05). There was no statistical significance (P>0.05) for the analyzed results for renal insufficiency or mortality. Of the 49 included studies, only 3 case studies of critically ill children reported deaths, and no deaths were reported in the remaining 47 studies. Publication bias was found for symptoms of sore throat, anhelation, headache, diarrhea, and vomiting, and no publication bias was found for the remaining clinical symptoms.

Among the 13 frequently reported laboratory findings (Table 4), the most common abnormalities in laboratory examinations were elevated procalcitonin (40% [0.23−0.57]), elevated lactate dehydrogenase (31% [0.19−0.43]), increased lymphocyte count (28% [0.17−0.42]), increased creatine kinase (28%[0.18−0.40]), and elevated C-reactive protein (26% [0.17−0.36]) (Supplementary Figs S 3, 4, 5, 6 and 7). Additional laboratory findings included elevated AST (24%[ 0.09−0.43]), decreased leukocyte count (21% [0.13−0.29]), elevated D-dimer (19% [0.09−0.31]), reduced lymphocyte count (15% [0.09−0.21]), increased leukocyte count (15% [0.07−0.25]), and elevated ALT (7% [0.05−0.10]). Because only two studies reported elevated creatinine, we did not perform a meta-analysis. All the results described above were statistically significant (p□<□0.05). Begg’s test indicated significant publication bias for decreased leukocyte count, elevated ALT, and elevated D-dimers, and no remaining laboratory findings showed significant publication bias at p < 0.05.

Among the 9 frequently reported chest CT findings (Table 5), in the included 26 studies, the prevalence of chest CT examinations showing infection was 67% [0.58−0.76] (Supplementary Figs S 8). The most common abnormalities from chest CT examination were lower lobe involvement (56% [0.42-0.70]), ground-glass opacities (33% [0.25−0.42]), bilateral pneumonia (32% [0.24-0.40]), patchy shadows (31% [0.18-0.45]), and upper lobe involvement (30% [0.20-0.41]) (Supplementary Figs S 9, 10, 11, 12 and 13). Other outcomes included unilateral pneumonia (26% [0.22-0.30]), tubercles (13% [0.06-0.21]), pleural fluid (3% [0.00-0.08]). All the abovementioned results were statistically significant (p□<□0.05). Publication bias was found for bilateral pneumonia, and there was no evidence of significant publication bias among the remaining 9 included studies.

## 3 Discussion

This meta-analysis summarizes the latest and most comprehensive study for clinical characteristics of COVID-19 in children and adolescents, from the first report of COVID-19 to July 10, 2020, including clinical features, laboratory outcomes, and chest CT findings. We conducted a systematic review and meta-analysis of 49 studies involving 1627 patients. The proportion of boys was significantly higher than that of girls, also consistent with previous studies, according to the synthesis results of our meta-analysis. Compared with the two previous systematic reviews on this topic, we have included more studies and analyzed 40 results in detail.

The main clinical features in children and adolescents with COVID-19 were fever (56%), cough (45%), sneezing (14%), vomiting (14%), and other less common results included diarrhea (12%), anhelation (11%), sore throat (10%), headache (10%), fatigue (9%), myalgia (8%), increased sputum volume (7%), and abdominal pain (6%). According to a previous study of COVID-19 in adult patients, the main clinical features of COVID-19 were fever, cough, and fatigue [59–60]. In this study of children and adolescents, patients with symptoms of sneezing and vomiting also accounted for reasonably large percentages. Previous studies have found that nasal congestion, runny nose and diarrhea were less common in children than in adults [61–63]. Critical or fatal pediatric COVID-19 cases were rare, and children with COVID-19 showed less severe clinical symptoms than did adults. In addition, one study showed that children may be less likely to be infected, and may show milder symptoms if they are infected [63]. Therefore, children and adolescents are more likely to have potential hidden infections, which should attract the attention of clinicians and parents of children.

The most common laboratory abnormalities were elevated procalcitonin (40%), elevated lactate dehydrogenase (31%), increased lymphocyte count (28%), elevated creatine kinase (28%) and elevated C-reactive protein (26%). These laboratory abnormalities indicate clear viral infection. Previous studies have found the most common laboratory test abnormalities in adult patients to be elevated C-reactive protein and reduced lymphocyte count [59–60]. All these abnormal laboratory markers are nonspecific, so their clinical applications are limited. According to the findings of our study, the elevated procalcitonin, elevated lactate dehydrogenase and increased lymphocyte count may appear as nonspecific markers in children and adolescents. Previous research has suggested that mortality, acute respiratory distress syndrome (ARDS), headache, increased leukocyte count, and elevated lactate dehydrogenase are significantly higher in studies with a larger proportion of older patients [59]. When evaluating suspicious cases, doctors cannot rely on these laboratory abnormalities to rule out or confirm the diagnosis of COVID-19 [59]. One study suggested that these abnormalities may be related to the cytokine storm caused by infection [64]. Recently, another study revealed that COVID-19 might mainly affect T lymphocytes, especially CD4+T cells, resulting in significant lymphopenia [65].The results of this study showed that the prevalence of increased lymphocyte count in children (28%) was higher than the prevalence of decreased lymphocyte count (15%), which may be related to the percentage of lymphocytes elevated in the growth and development of the child’s immune system. In terms of liver enzymes, elevated AST (24%) was more common than elevated ALT (7%), which indicates that liver function may be damaged to varying degrees. In addition, reduced leukocyte count (21%) was more common than increased leukocyte count (15%). Of the 49 studies reviewed, only two reported elevated creatinine. Therefore, elevated creatinine was extremely rare among children with COVID-19.

Chest CT examination was one of the important auxiliary examinations for COVID-19 pneumonia, and it has a very satisfactory effectiveness for the examination of lung lesions. Previous research has found that most patients have mild symptoms and elevated body temperature, but pulmonary lesions are relatively obvious. Li [66] found that chest CT had a low rate of missed diagnosis of COVID-19 (3.9%) and could thus be used as a rapid diagnostic method for this disease to optimize patient management. However, CT is still limited in identifying specific viruses and distinguishing viruses. Our meta-analysis showed that the prevalence of children and adolescents with COVID-19 chest CT examinations showing infection was 67%. A recent study of 62 adult patients with COVID-19 showed that 52 (83.9%) had an abnormal chest CT scan, which was much higher than the frequency in children and adolescents with an abnormal chest CT scan, consistent with previous reports [67]. The results of our meta-analysis showed that the prevalence of ground-glass opacity and patchy shadows were 33% and 31%, respectively. Lung lesions of patients with COVID-19 showed imageology findings similar to those of other viral pneumonia, mainly ground-glass opacities and patchy shadows. The most common position for lesions detected by CT scan was in the lower lobe (56%), while incidence of upper lobe lesion was 30%. Bilateral pneumonia (32%) was more common than unilateral pneumonia (26%). A study of adult COVID-19 patients showed that bilateral pneumonia was present in 73.2% of cases, indicating that adults are more likely to experience bilateral lung infection than are children and adolescents [59]. The results of this study found that tubercles (13%) and pleural fluid (3%) rarely occur in children and adolescents with COVID-19. Chang et al. found that, compared with SARS, COVID-19 has prominent characteristics in children, and the typical appearance of chest CT is ground-glass opacity, which is also consistent with our study [68]. Other CT findings, such as air bronchogram and lymphadenopathy, are less common [69].

Among the 49 articles included, only 3 studies of critically ill children reported deaths. Among the 1627 patients included, only 7 deaths occurred, and the overall mortality was lower than the reported adult mortality (3.6%) [59]. Compared with other common viral diseases, the number of cases of COVID-19 in children and adolescents is small, and the mortality is low, which is very gratifying.

There are also several limitations to our systematic review and meta-analysis. First, all reported cases were retrospective studies, and there was a high degree of heterogeneity among the results of the study. Because there were many studies included, it is difficult to avoid heterogeneity. No subgroup analysis was performed on different age groups of the included children and adolescents. Second, the current COVID-19 pandemic has not been effectively controlled. Our study was analyzed during the COVID-19 outbreak. Most of the included patients were from China, and there may be certain geographical and ethnic differences. Subgroup analysis was not performed, which may also bias the results of the analysis.

## 4 Conclusion

This meta-analysis provides a comprehensive review of the clinical characteristics of COVID-19 in children and adolescents. The main clinical features of children and adolescents were fever and cough. The most frequently reported laboratory abnormalities were elevated procalcitonin, elevated lactate dehydrogenase, increased lymphocyte count, increased creatine kinase, and elevated C-reactive protein. From chest computed tomography, the most common lesion site was the lower lobe. Bilateral pneumonia was more common than unilateral pneumonia, and the main manifestations were ground-glass opacities and patchy shadowing. The condition of children and adolescents with COVID-19 was milder than that of adult patients, with a greater proportion of asymptomatic and mild cases, and specific diagnosis and source-of-infection control were more challenging.

## Data Availability

All data generated or analyzed during this study are included in this article.

## Declarations

### Ethics approval and consent to participate

Not applicable. This article does not contain any studies with human participants performed by any of the authors.

## Consent for publication

All participating authors provide consent for publication.

## Availability of data and materials

All data generated or analyzed during this study are included in this article.

## Competing interests

The authors declare that they have no competing interests.

## Funding

The authors state that this work has not received financial support.

## Authors’ contributions

All the authors designed the study. LL, HZ, ZL and BT designed the literature search and searched the articles. LL, HZ, ML, YC, JL, and BT contributed to statistical analysis, interpretation of data and data review. LL and HZ wrote the first draft of the article. All the authors revised the article and approved the final version.

## Acknowledgements

Not applicable.

---

**Figure S 1.**
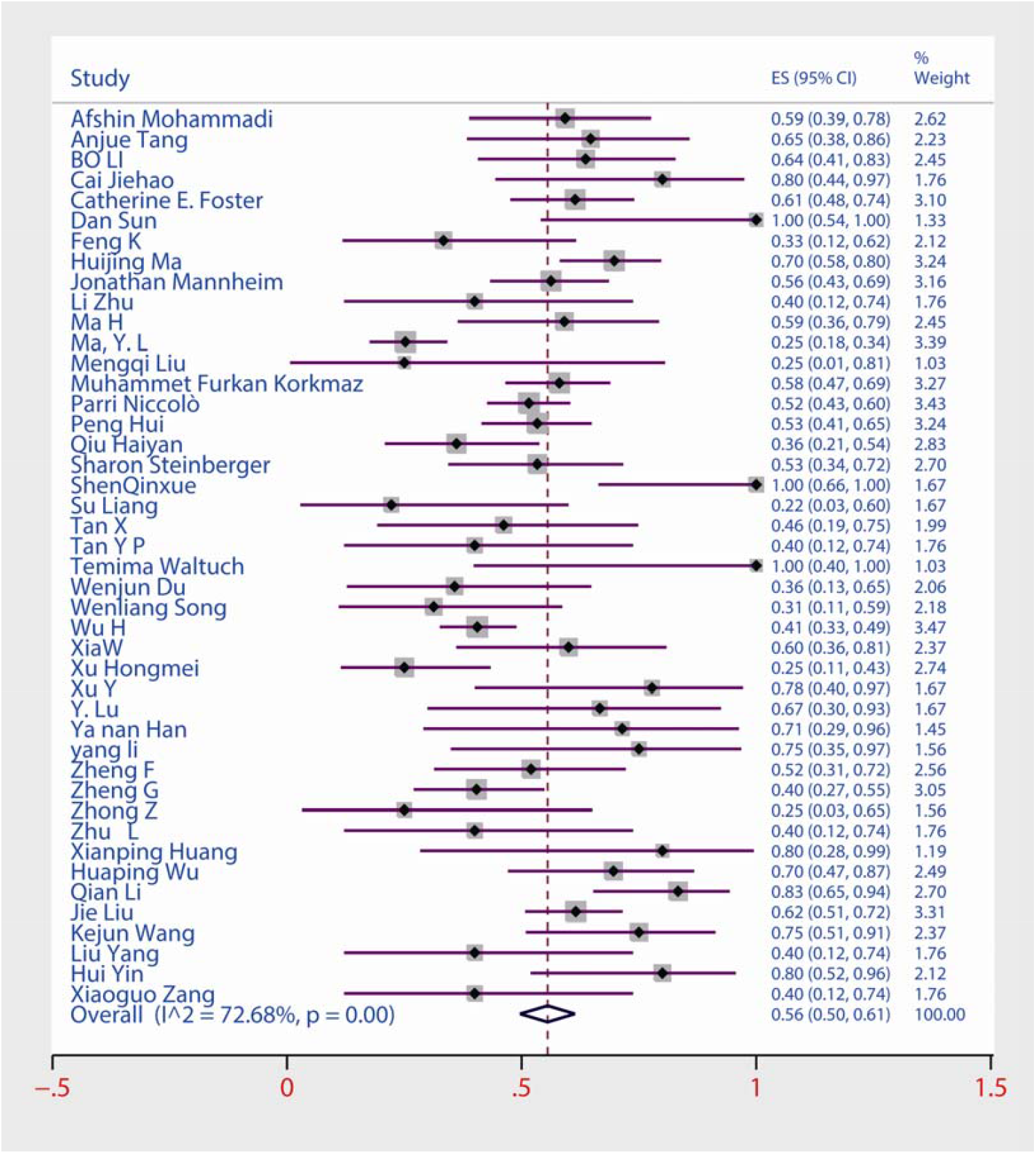
Forest plots of the prevalence of fever among children and adolescents with COVID-19.

**Figure S 2.**
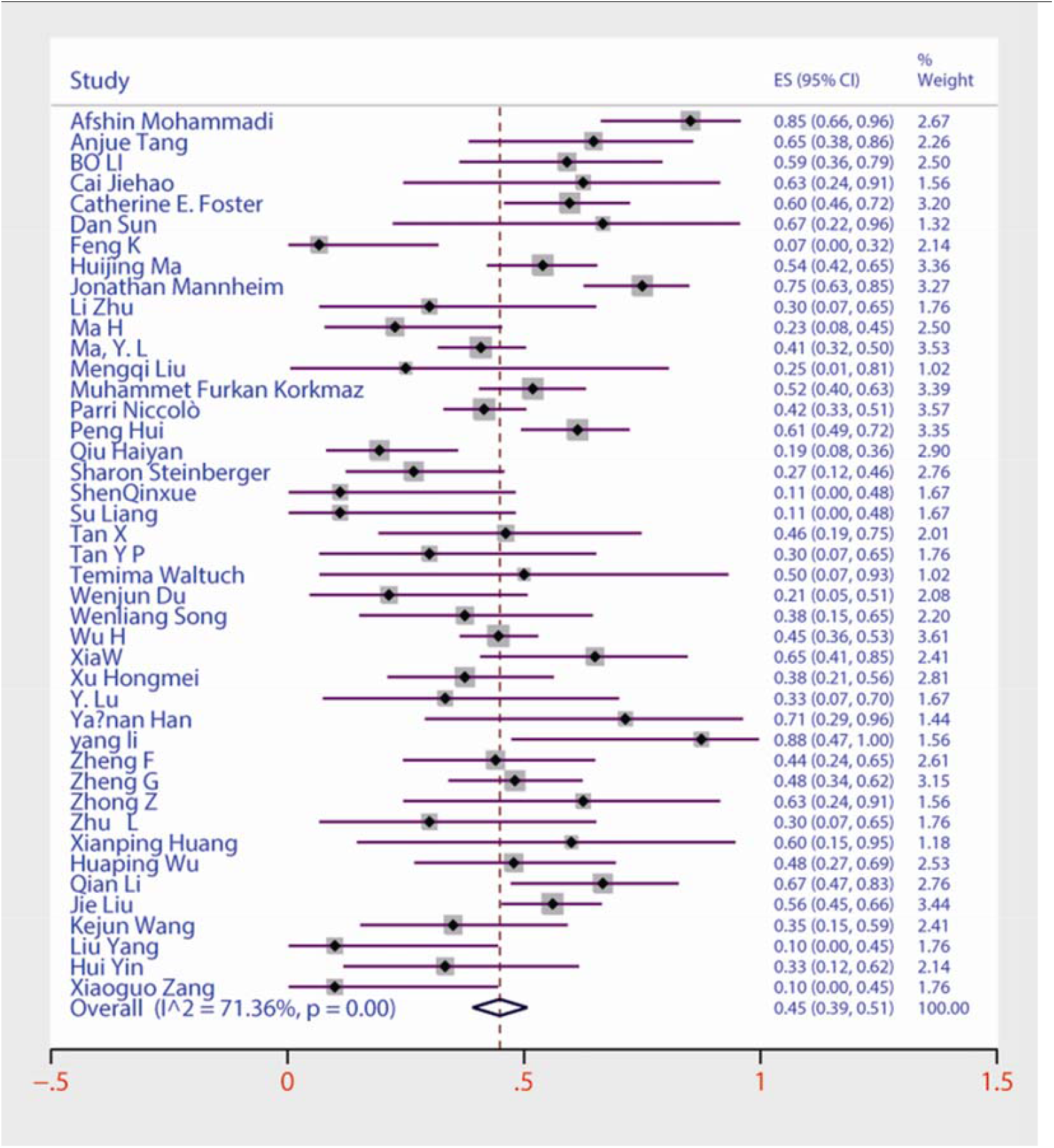
Forest plots of the prevalence of cough among children and adolescents with COVID-19.

**Figure S 3.**
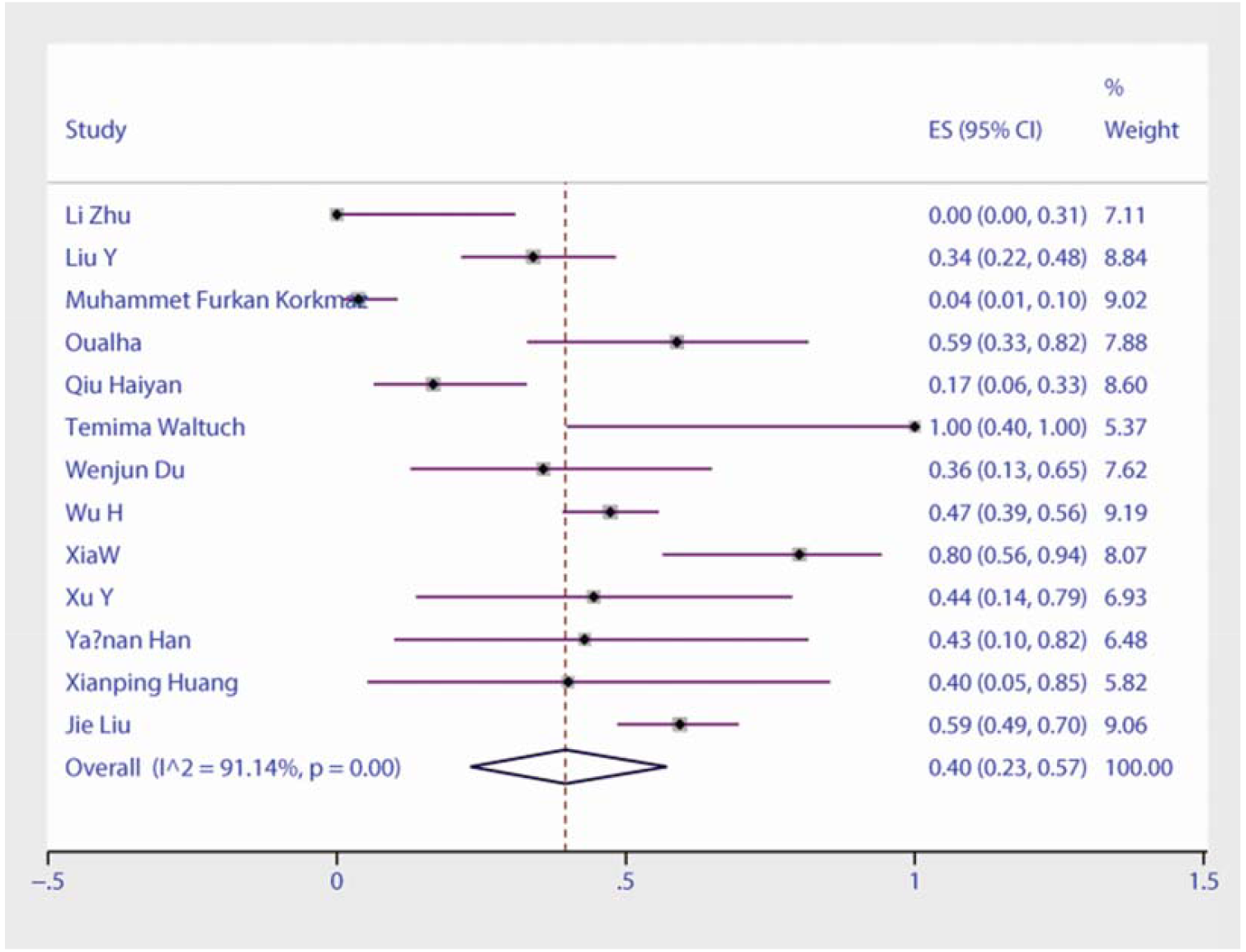
Forest plots of the prevalence of elevated procalcitonin among children and adolescents with COVID-19.

**Figure S 4.**
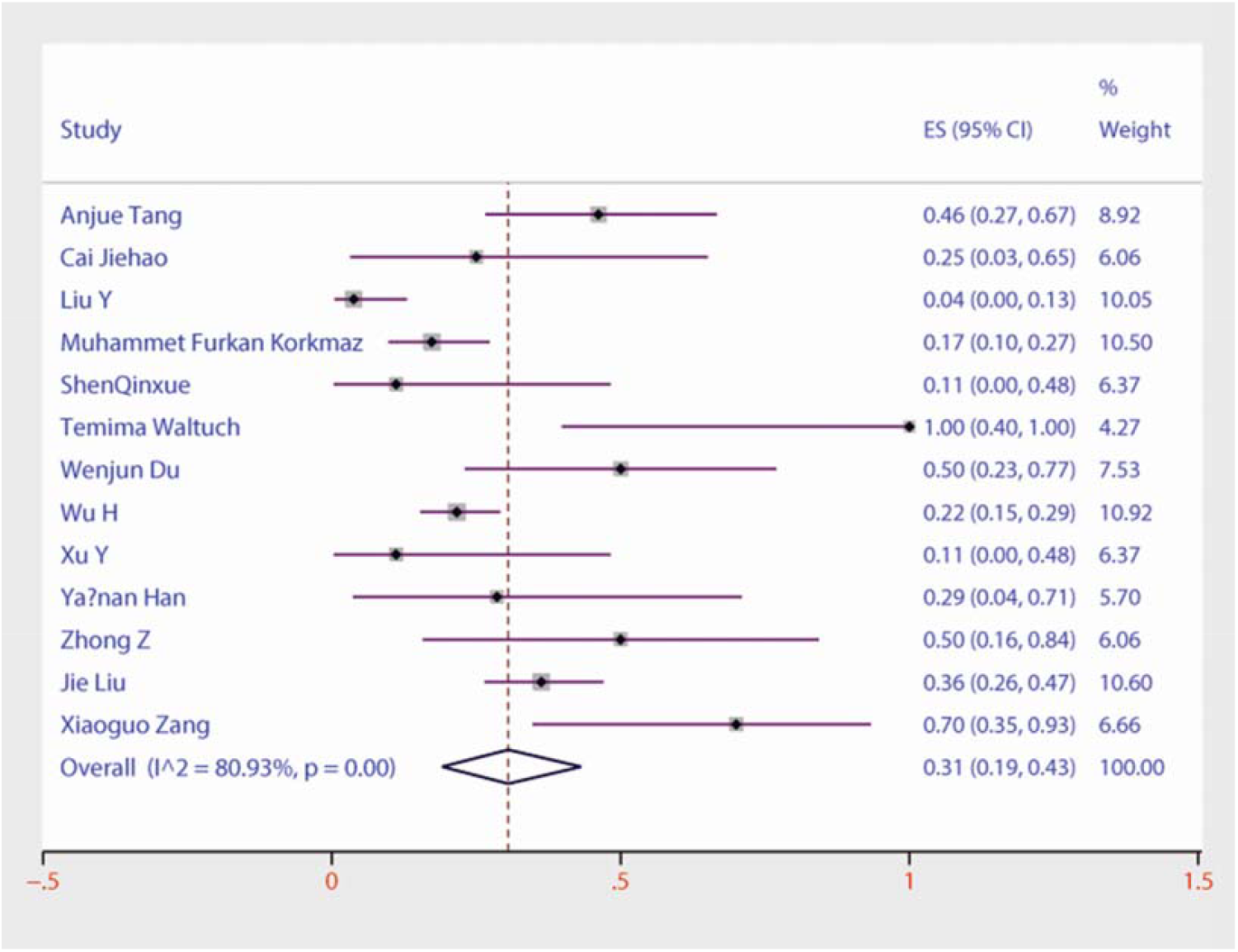
Forest plots of the prevalence of elevated lactate dehydrogenase among children and adolescents with COVID-19.

**Figure S 5.**
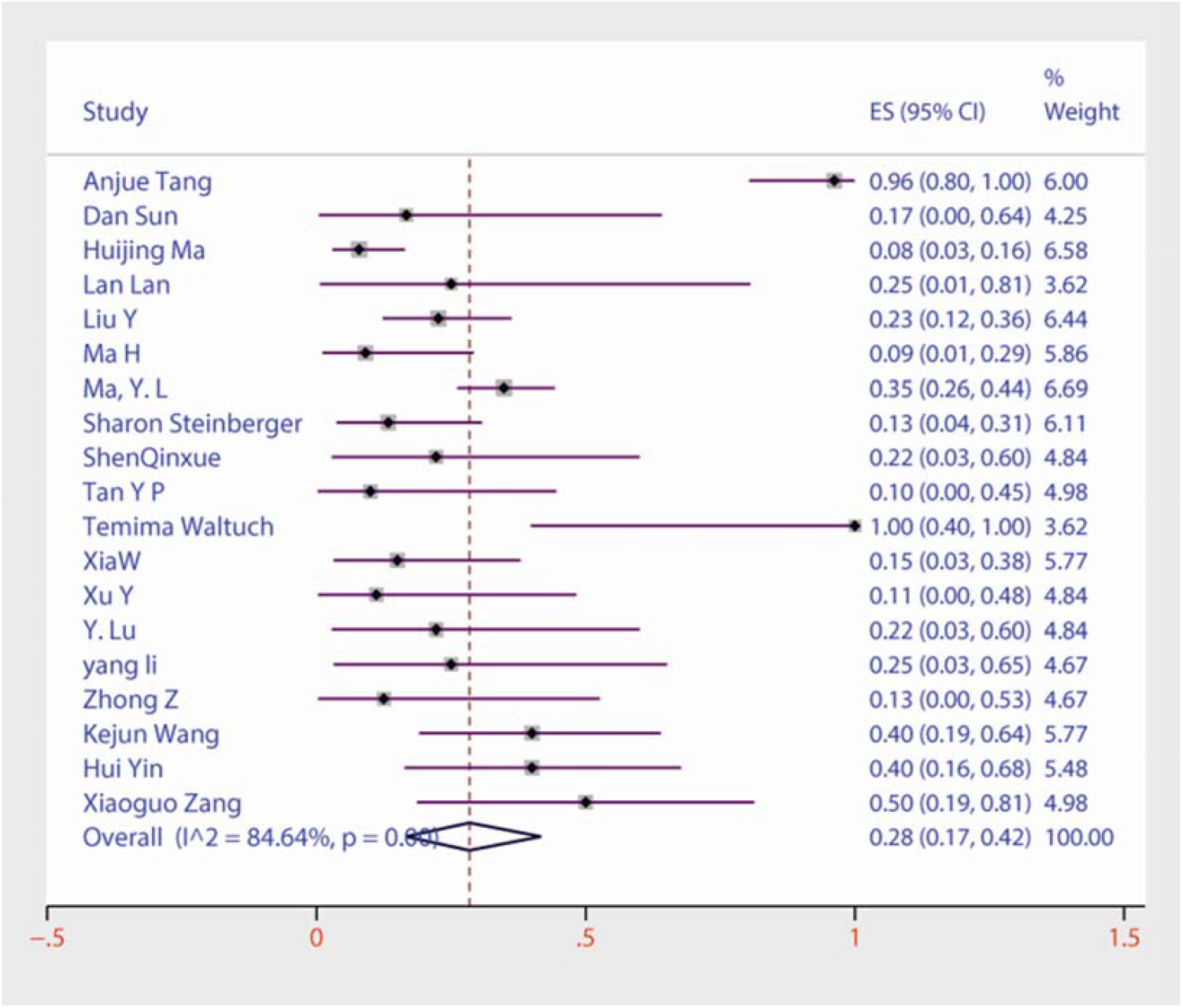
Forest plots of the prevalence of increased lymphocyte count among children and adolescents with COVID-19.

**Figure S 6.**
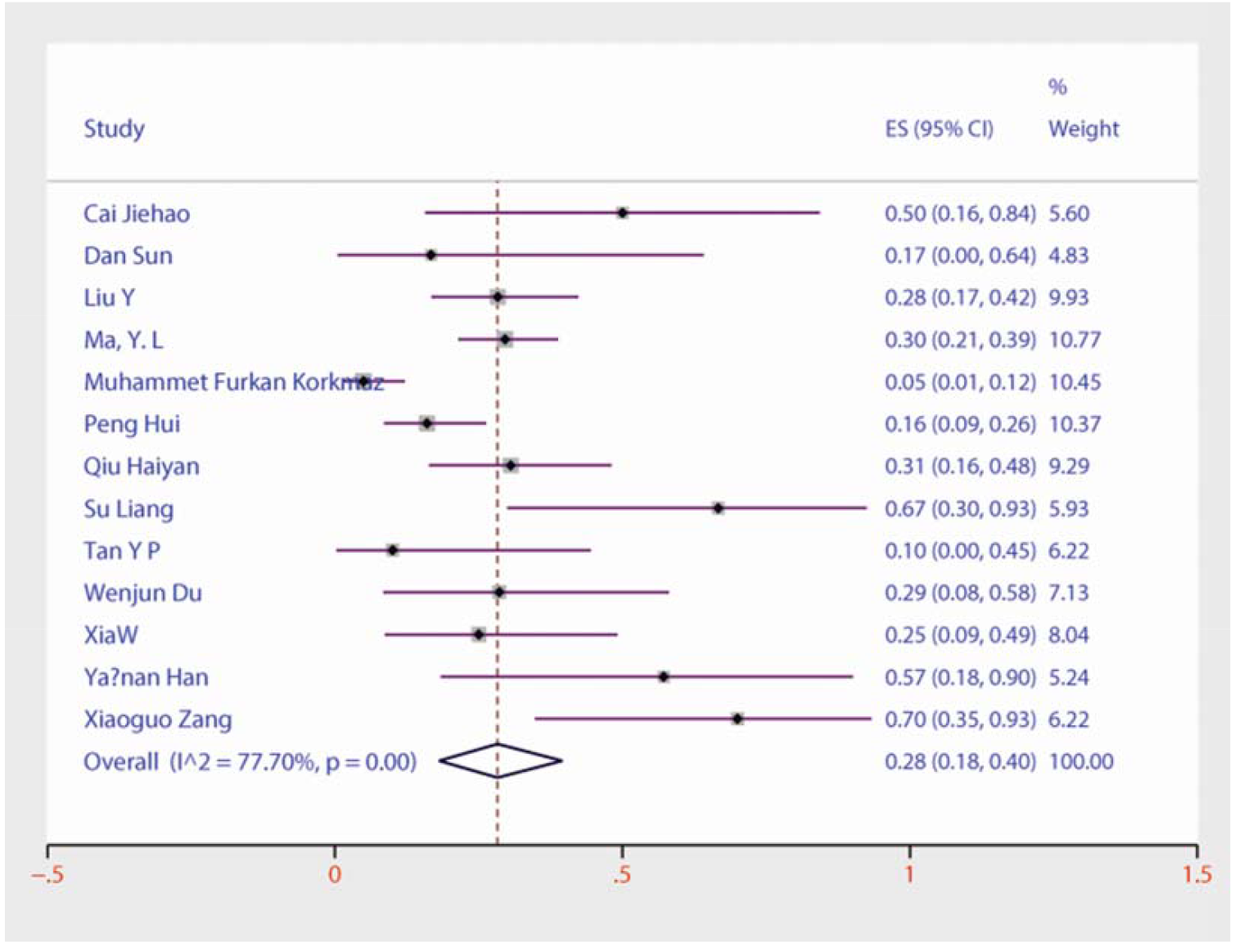
Forest plots of the prevalence of increased creatine kinase among children and adolescents with COVID-19.

**Figure S 7.**
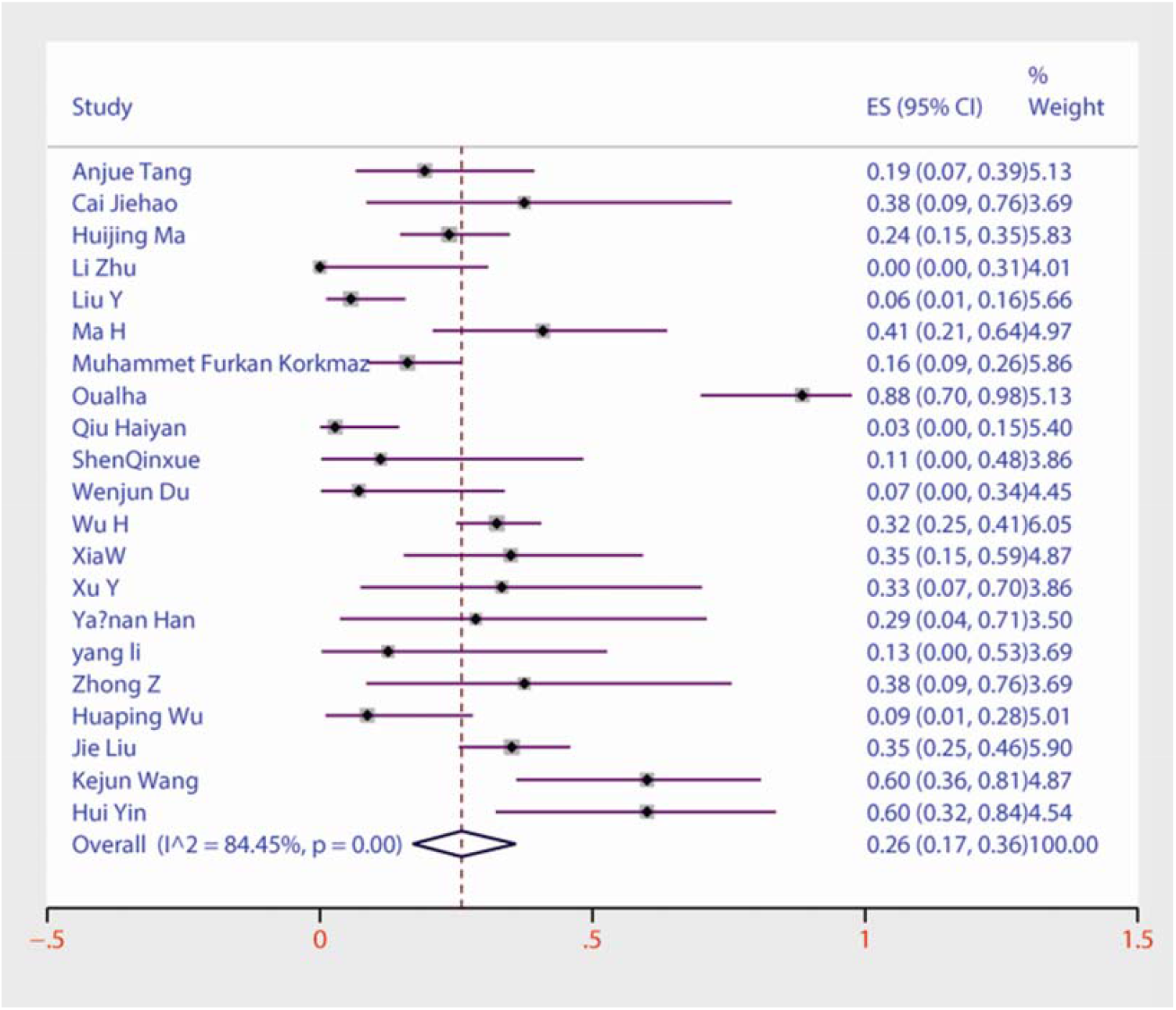
Forest plots of the prevalence of C-reactive protein among children and adolescents with COVID-19.

**Figure S 8.**
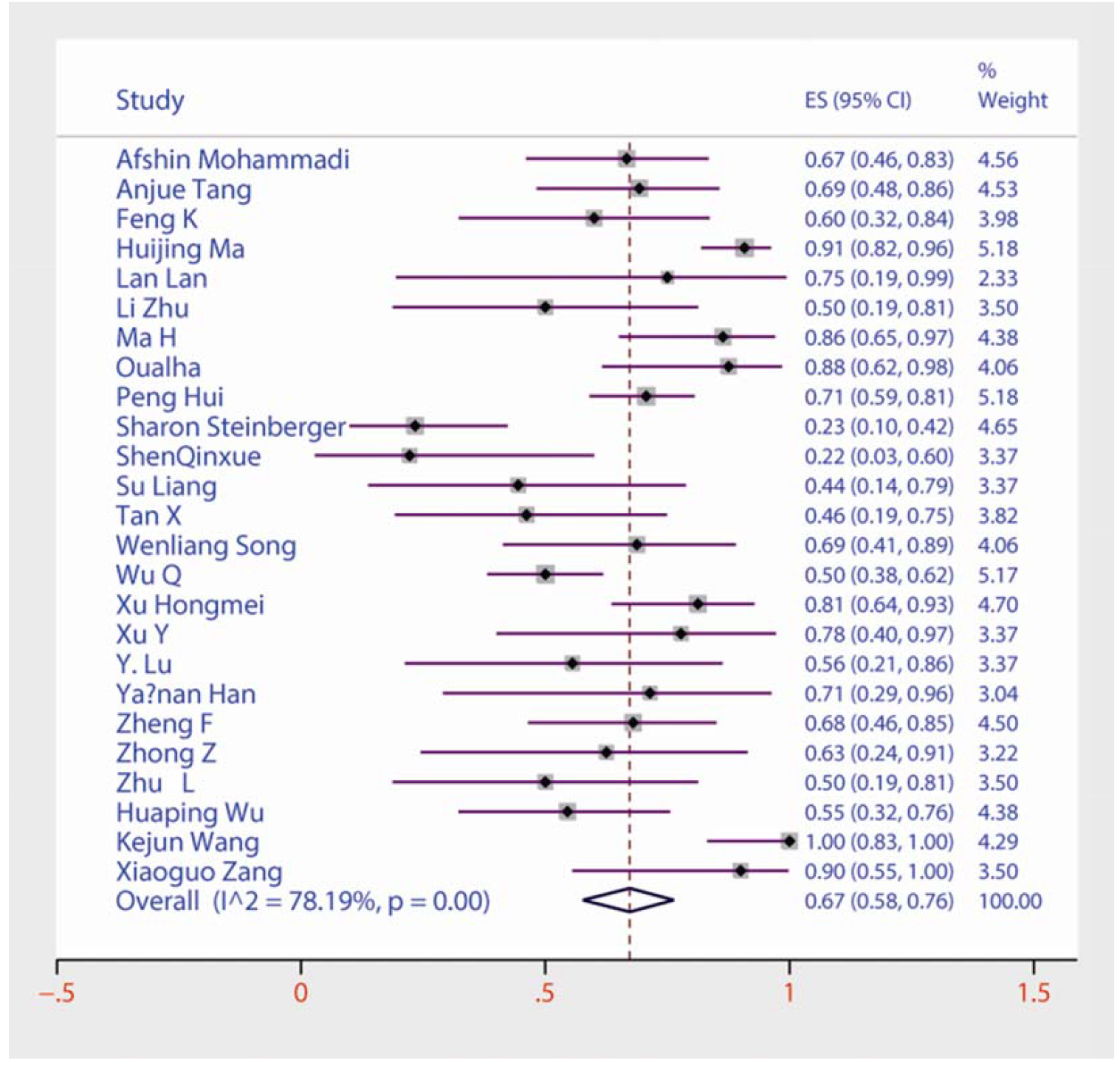
Forest plots of the prevalence of chest CT examinations showing infection among children and adolescents with COVID-19.

**Figure S 9.**
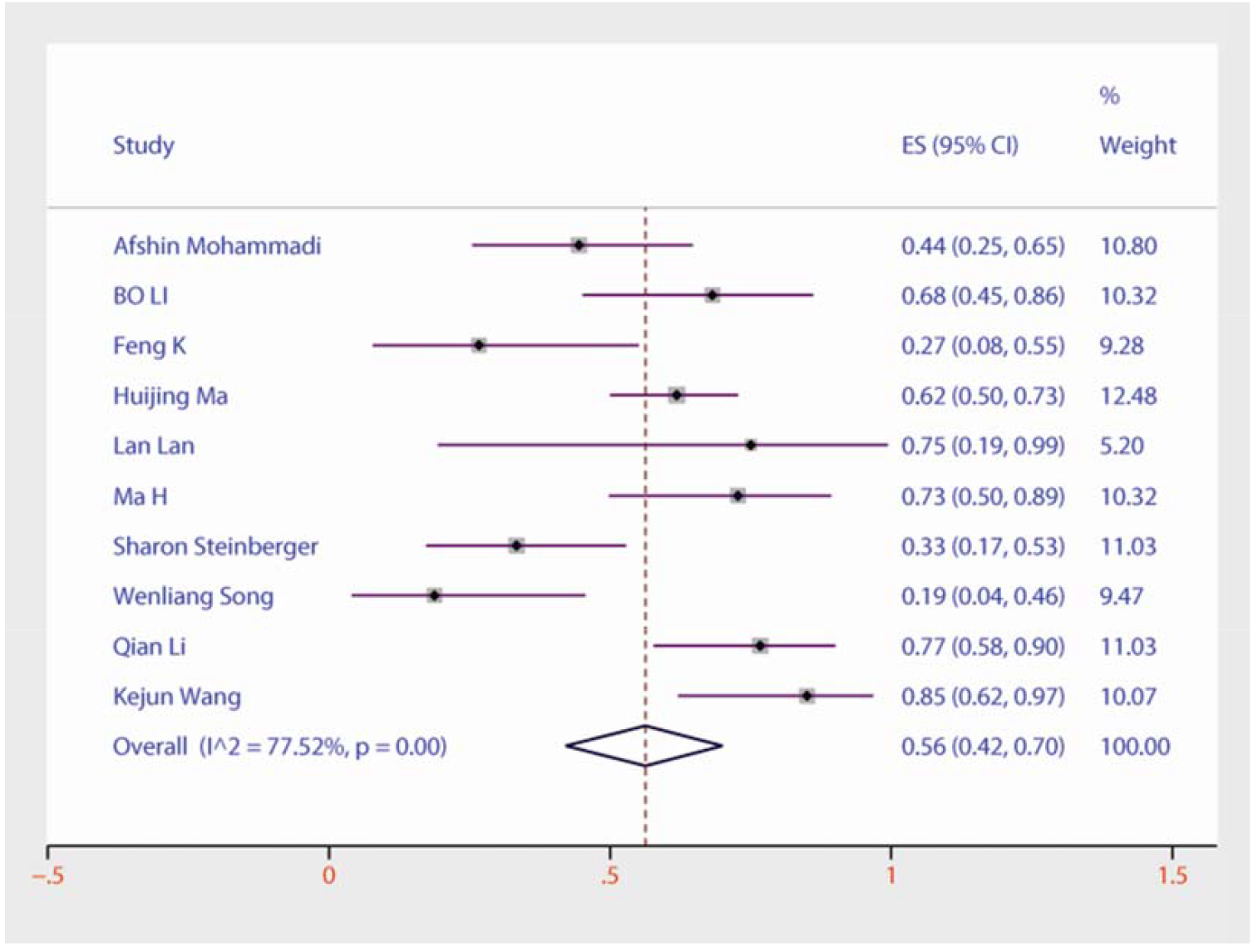
Forest plots of the prevalence of lower lobe involvement among children and adolescents with COVID-19.

**Figure S 10.**
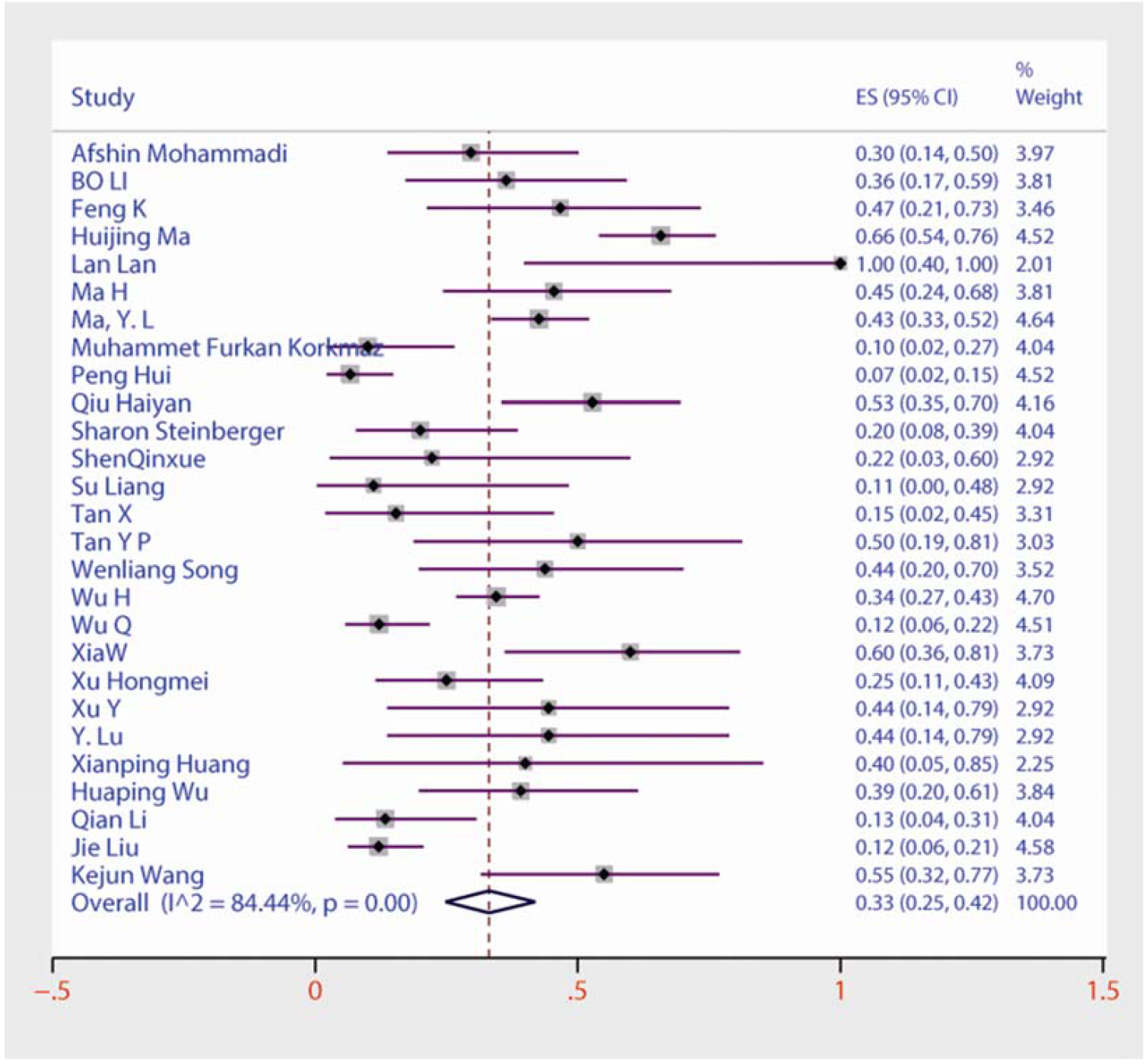
Forest plots of the prevalence of ground-glass opacities among children and adolescents with COVID-19.

**Figure S 11.**
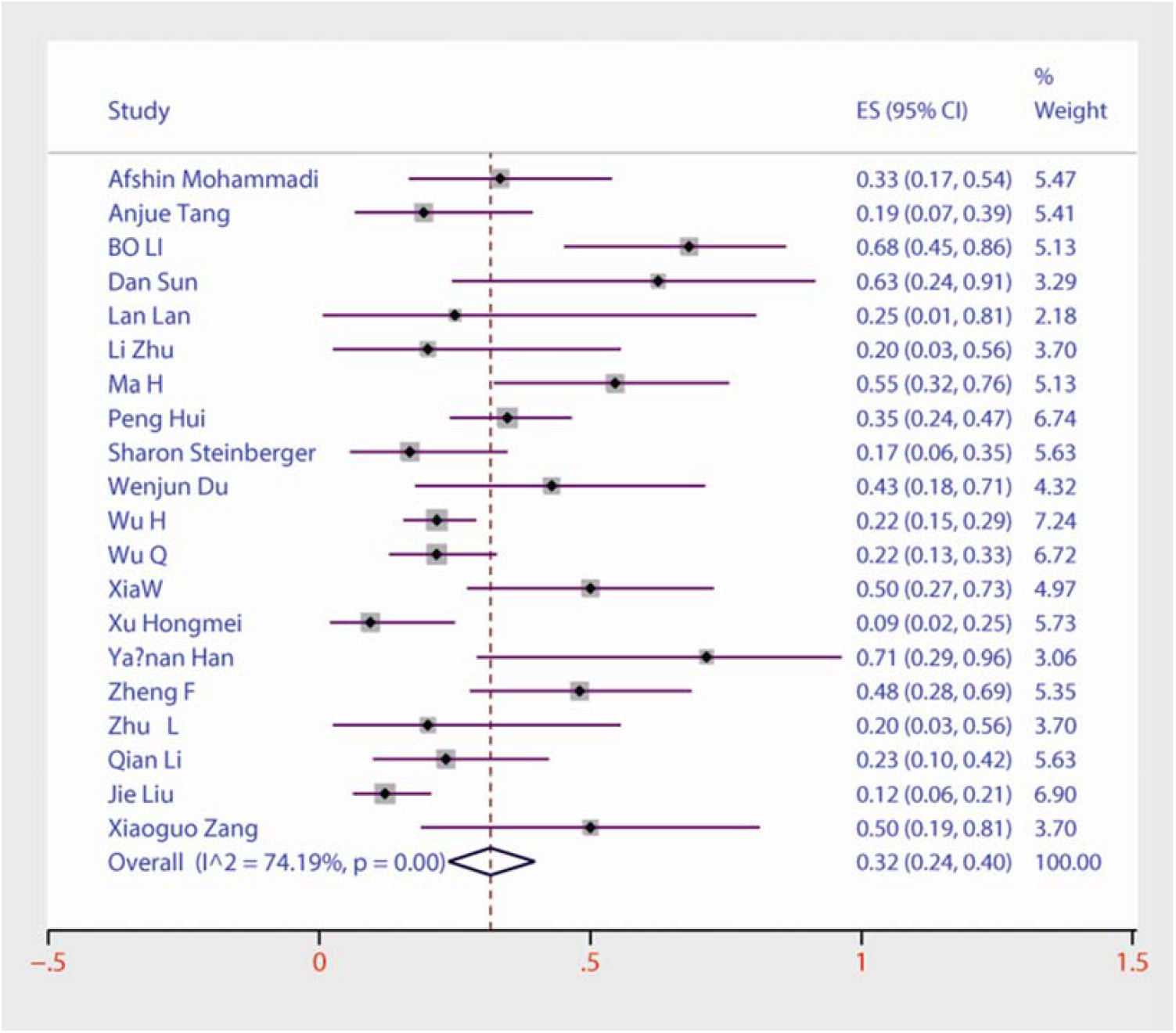
Forest plots of the prevalence of bilateral pneumonia among children and adolescents with COVID-19.

**Figure S 12.**
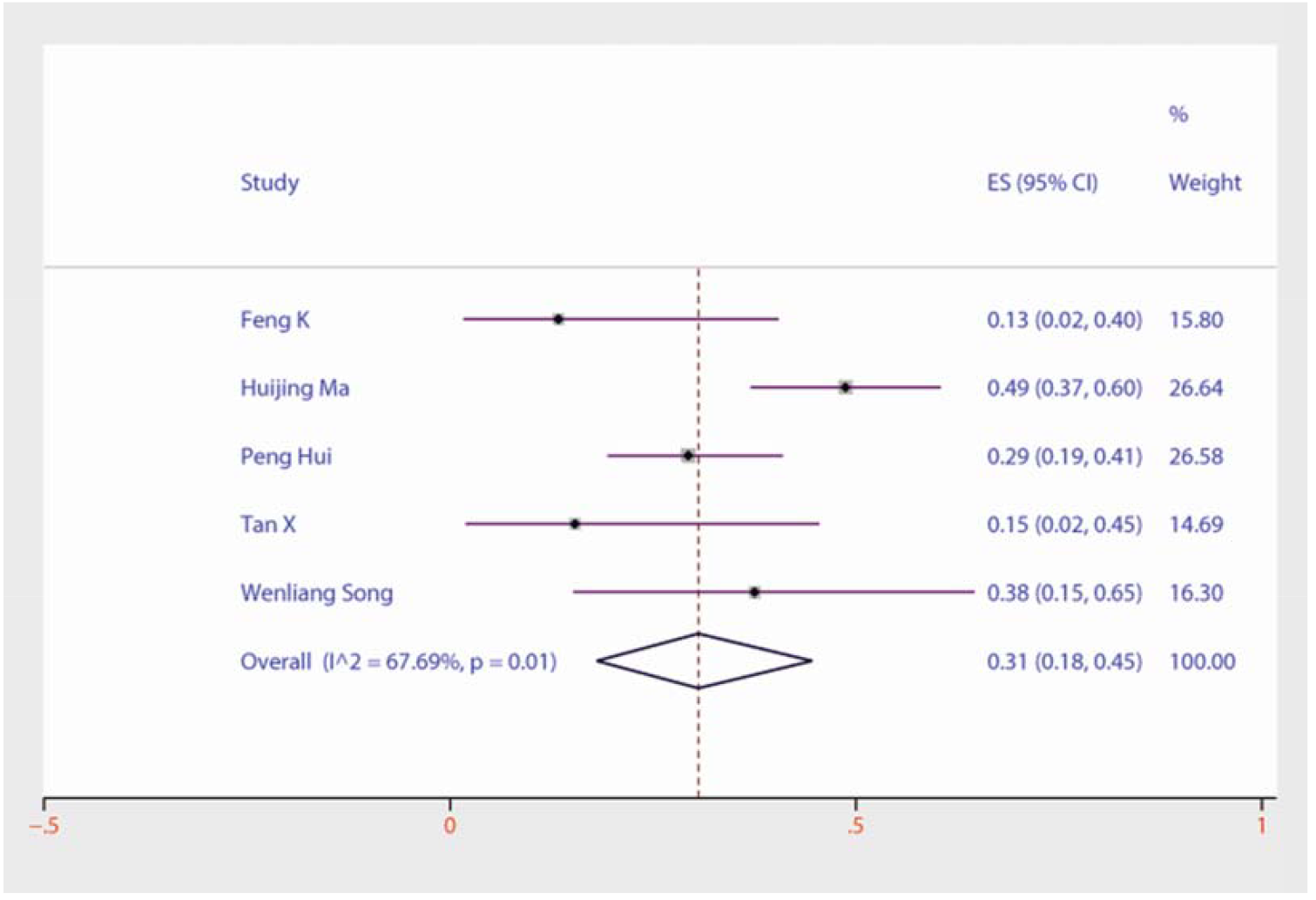
Forest plots of the prevalence of patchy shadows among children and adolescents with COVID-19.

**Figure S 13.**
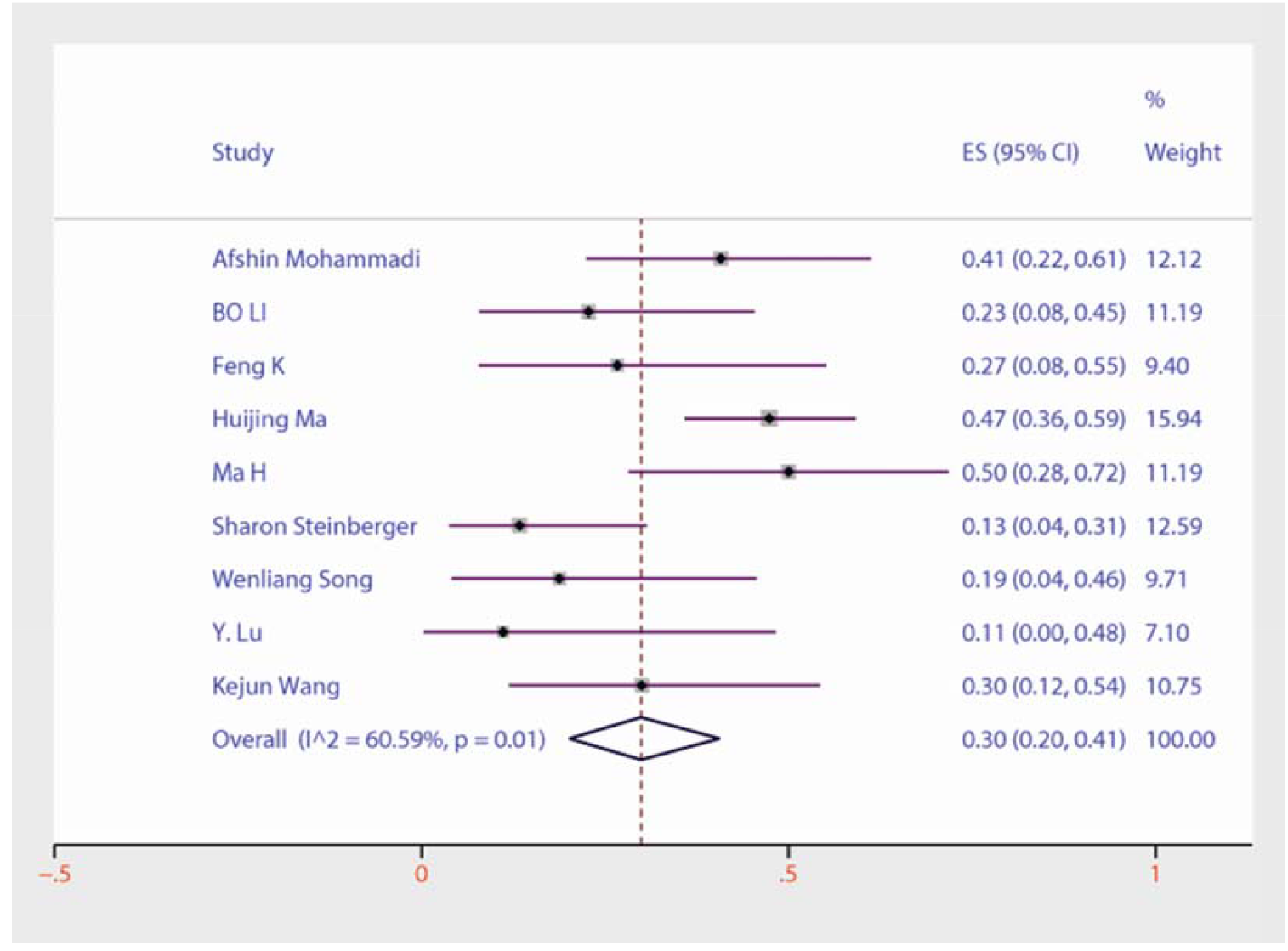
Forest plots of the prevalence of upper lobe involvement among children and adolescents with COVID-19.

## Notes

### Competing Interest Statement

The authors have declared no competing interest.

## References

[1] Wang C, Horby PW, Hayden FG, Gao GF. A novel coronavirus outbreak of global health concern. Lancet. 2020;395(10223):470–473. doi: 10.1016/S0140-6736(20)30185-9.

[2] World Health Organization. Clinica lmanagement of severe acute respiratory infection when when novelcoronavirus(nCoV) infection issuspected:interimguidance,25January2020[R].WHO,2020.

[3] Hoffmann M, Kleine-Weber H, Schroeder S, Krüger N, Herrler T, Erichsen S, Schiergens TS, Herrler G, Wu NH, Nitsche A, Müller MA. SARS-CoV-2 cell entry depends on ACE2 and TMPRSS2 and is blocked by a clinically proven proteaseinhibitor. Cell. 2020 Mar 5.

[4] van Doremalen, Neeltje, Trenton Bushmaker, Dylan H. Morris, Myndi G. Holbrook,Amandine Gamble, Brandi N. Williamson, Azaibi Tamin et al. “Aerosol and surfacestability of SARS-CoV-2 as compared with SARS-CoV-1.” New England Journal of Medicine (2020).

[5] Xu, Yi, Xufang Li, Bing Zhu, Huiying Liang, Chunxiao Fang, Yu Gong, Qiaozhi Guo et al. “Characteristics of pediatric SARS-CoV-2 infection and potential evidence for persistent fecal viral shedding.” Nature Medicine (2020): 1–4.doi:10.1038/s41591-020-0817-4.

[6] YangY LQ, Liu M, et al. Epidemiological and clinical features of the 2019 novel coronavirus outbreak in China. medRxiv preprint 2020

[7] Liberati A, Altman DG, Tetzlaff J, et al. The PRISMA statement for reporting systematic reviews and meta-analyses of studies that evaluate healthcare interventions: explanation and elaboration. BMJ. 2009;339:b2700. Published 2009 Jul 21. doi:10.1136/bmj.b2700

[8] Stroup DF, Berlin JA, Morton SC, et al. Meta-analysis of observational studies in epidemiology: a proposal for reporting. Meta-analysis Of Observational Studies in Epidemiology (MOOSE) group. JAMA. 2000;283(15):2008–2012. doi:10.1001/jama.283.15.2008

[9] American National Institute of Health. https://www.nhlbi.nih.gov/health-topics/study-quality-assessment-tools.

[10] Mohammadi A, Mohebbi I, Khademvatani K, et al. Clinical and radiological characteristics of pediatric patients with COVID-19: focus on imaging findings [published online ahead of print, 2020 Jun 13]. Jpn J Radiol. 2020;1–6. doi:10.1007/s11604-020-01003-6

[11] Tang A, Xu W, Chen P, et al. A retrospective study of the clinical characteristics of COVID-19 infection in 26 children[J]. medRxiv, 2020. doi: 10.1101/2020.03.08.20029710

[12] Li B, Shen J, Li L, Yu C. Radiographic and Clinical Features of Children With Coronavirus Disease (COVID-19) Pneumonia. Indian Pediatr. 2020;57(5):423–426. doi:10.1007/s13312-020-1816-8

[13] Cai J, Xu J, Lin D, et al. A Case Series of children with 2019 novel coronavirus infection: clinical and epidemiological features [published online ahead of print, 2020 Feb 28]. Clin Infect Dis. 2020;ciaa198. doi:10.1093/cid/ciaa198

[14] Foster CE, Moulton EA, Munoz FM, et al. Coronavirus Disease 2019 in Children Cared for at Texas Children’s Hospital: Initial Clinical Characteristics and Outcomes. J Pediatric Infect Dis Soc. 2020;9(3):373–377. doi:10.1093/jpids/piaa072

[15] Sun D, Li H, Lu XX, et al. Clinical features of severe pediatric patients with coronavirus disease 2019 in Wuhan: a single center’s observational study. World J Pediatr. 2020;16(3):251–259. doi:10.1007/s12519-020-00354-4

[16] Feng K, Yun Y X, Wang X F, et al. Analysis of CT features of 15 children with 2019 novel coronavirus infection. Zhonghua er ke za zhi= Chinese journal of pediatrics, 2020, 58: E007–E007. doi: 10.3760/cma.j.cn112140-20200210-00071

[17] Ma H, Hu J, Tian J, et al. A single-center, retrospective study of COVID-19 features in children: a descriptive investigation. BMC Med. 2020;18(1):123. Published 2020 May 6. doi:10.1186/s12916-020-01596-9

[18] Mannheim J, Gretsch S, Layden JE, Fricchione MJ. Characteristics of Hospitalized Pediatric COVID-19 Cases - Chicago, Illinois, March - April 2020 [published online ahead of print, 2020 Jun 1]. J Pediatric Infect Dis Soc. 2020;piaa070. doi:10.1093/jpids/piaa070

[19] Lan L, Xu D, Xia C, Wang S, Yu M, Xu H. Early CT Findings of Coronavirus Disease 2019 (COVID-19) in Asymptomatic Children: A Single-Center Experience. Korean J Radiol. 2020;21(7):919–924. doi:10.3348/kjr.2020.0231

[20] Zhu L, Wang J, Huang R, et al. Clinical characteristics of a case series of children with coronavirus disease 2019. Pediatr Pulmonol. 2020;55(6):1430–1432. doi:10.1002/ppul.24767

[21] Liu YJ, Chen P, Liu ZS, et al. Clinical features of asymptomatic or subclinical COVID-19 in children. Chinese Journal of Contemporary Pediatrics, 2020, 22(6): 578. doi:10.7499/j.issn.1008-8830.2004088 (in chinese)

[22] Ma H, Shao J, Wang Y, et al. High resolution CT features of COVID-19 in children. Chinese Journal of Radiology (China), 2020, 54(4) : 310–313. doi: 10.3760/cma.j.cn112149-20200206-00100

[23] Ma YL, Xia SY, Wang M, et al. Clinical features of children with SARS-CoV-2 infection: an analysis of 115 cases. Chinese Journal of Contemporary Pediatrics, 2020, 22(4): 290–293. doi:10.7499/j.issn.1008-8830.2003016 (in chinese)

[24] Liu M, Song Z, Xiao K. High-Resolution Computed Tomography Manifestations of 5 Pediatric Patients With 2019 Novel Coronavirus. J Comput Assist Tomogr. 2020;44(3):311–313. doi:10.1097/RCT.0000000000001023

[25] Korkmaz MF, Türe E, Dorum BA, Kılıç ZB. The Epidemiological and Clinical Characteristics of 81 Children with COVID-19 in a Pandemic Hospital in Turkey: an Observational Cohort Study. J Korean Med Sci. 2020;35(25):e236. Published 2020 Jun 29. doi:10.3346/jkms.2020.35.e236

[26] Oualha M, Bendavid M, Berteloot L, et al. Severe and fatal forms of COVID-19 in children. Arch Pediatr. 2020;27(5):235–238. doi:10.1016/j.arcped.2020.05.010

[27] Parri N, Magistà AM, Marchetti F, et al. Characteristic of COVID-19 infection in pediatric patients: early findings from two Italian Pediatric Research Networks. Eur J Pediatr. 2020;179(8):1315–1323. doi:10.1007/s00431-020-03683-8

[28] Peng H, Gao P, Xu Q, et al. Coronavirus disease 2019 in children: Characteristics, antimicrobial treatment, and outcomes. J Clin Virol. 2020;128:104425. doi:10.1016/j.jcv.2020.104425

[29] Qiu H, Wu J, Hong L, Luo Y, Song Q, Chen D. Clinical and epidemiological features of 36 children with coronavirus disease 2019 (COVID-19) in Zhejiang, China: an observational cohort study. Lancet Infect Dis. 2020;20(6):689–696. doi:10.1016/S1473-3099(20)30198-5

[30] Steinberger S, Lin B, Bernheim A, et al. CT Features of Coronavirus Disease (COVID-19) in 30 Pediatric Patients [published online ahead of print, 2020 May 22]. AJR Am J Roentgenol. 2020;1–9. doi:10.2214/AJR.20.23145

[31] Shekerdemian LS, Mahmood NR, Wolfe KK, et al. Characteristics and Outcomes of Children With Coronavirus Disease 2019 (COVID-19) Infection Admitted to US and Canadian Pediatric Intensive Care Units [published online ahead of print, 2020 May 11]. JAMA Pediatr. 2020;10.1001/jamapediatrics.2020.1948. doi:10.1001/jamapediatrics.2020.1948

[32] Shen Q, Guo W, Guo T, et al. Novel coronavirus infection in children outside of Wuhan, China. Pediatr Pulmonol. 2020;55(6):1424–1429. doi:10.1002/ppul.24762

[33] Su L, Ma X, Yu H, et al. The different clinical characteristics of corona virus disease cases between children and their families in China - the character of children with COVID-19. Emerg Microbes Infect. 2020;9(1):707–713. doi:10.1080/22221751.2020.1744483

[34] Tan X, Huang J, Zhao F, et al. Clinical features of children with SARS-CoV-2 infection: an analysis of 13 cases from Changsha, China. Zhongguo dang dai er ke za zhi= Chinese journal of contemporary pediatrics, 2020, 22(4): 294. doi:10.7499/j.issn.1008-8830.2003199

[35] Tan YP, Tan BY, Pan J, Wu J, Zeng SZ, Wei HY. Epidemiologic and clinical characteristics of 10 children with coronavirus disease 2019 in Changsha, China. J Clin Virol. 2020;127:104353. doi:10.1016/j.jcv.2020.104353

[36] Waltuch T, Gill P, Zinns LE, et al. Features of COVID-19 post-infectious cytokine release syndrome in children presenting to the emergency department [published online ahead of print, 2020 May 23]. Am J Emerg Med. 2020;S0735-6757(20)30403-4. doi:10.1016/j.ajem.2020.05.058

[37] Du W, Yu J, Wang H, et al. Clinical characteristics of COVID-19 in children compared with adults in Shandong Province, China. Infection. 2020;48(3):445–452. doi:10.1007/s15010-020-01427-2

[38] Song W, Li J, Zou N, Guan W, Pan J, Xu W. Clinical features of pediatric patients with coronavirus disease (COVID-19). J Clin Virol. 2020;127:104377. doi:10.1016/j.jcv.2020.104377

[39] Wu H, Zhu H, Yuan C, et al. Clinical and Immune Features of Hospitalized Pediatric Patients With Coronavirus Disease 2019 (COVID-19) in Wuhan, China. JAMA Netw Open. 2020;3(6):e2010895. Published 2020 Jun 1. doi:10.1001/jamanetworkopen.2020.10895

[40] Wu Q, Xing Y, Shi L, et al. Coinfection and Other Clinical Characteristics of COVID-19 in Children. Pediatrics. 2020;146(1):e20200961. doi:10.1542/peds.2020-0961

[41] Xia W, Shao J, Guo Y, Peng X, Li Z, Hu D. Clinical and CT features in pediatric patients with COVID-19 infection: Different points from adults. Pediatr Pulmonol. 2020;55(5):1169–1174. doi:10.1002/ppul.24718

[42] Xu H, Liu E, Xie J, et al. A follow-up study of children infected with SARS-CoV-2 from western China. Ann Transl Med. 2020;8(10):623. doi:10.21037/atm-20-3192

[43] Xu H, Liu E, Xie J, et al. A follow-up study of children infected with SARS-CoV-2 from western China. Ann Transl Med. 2020;8(10):623. doi:10.21037/atm-20-3192

[44] Lu Y, Wen H, Rong D, Zhou Z, Liu H. Clinical characteristics and radiological features of children infected with the 2019 novel coronavirus. Clin Radiol. 2020;75(7):520–525. doi:10.1016/j.crad.2020.04.010

[45] Han YN, Feng ZW, Sun LN, et al. A comparative-descriptive analysis of clinical characteristics in 2019-coronavirus-infected children and adults [published online ahead of print, 2020 Apr 6]. J Med Virol. 2020;10.1002/jmv.25835. doi:10.1002/jmv.25835

[46] Li Y, Cao J, Zhang X, Liu G, Wu X, Wu B. Chest CT imaging characteristics of COVID-19 pneumonia in preschool children: a retrospective study. BMC Pediatr. 2020;20(1):227. Published 2020 May 18. doi:10.1186/s12887-020-02140-7

[47] Zheng F, Liao C, Fan QH, et al. Clinical Characteristics of Children with Coronavirus Disease 2019 in Hubei, China. Curr Med Sci. 2020;40(2):275–280. doi:10.1007/s11596-020-2172-6

[48] Zheng G, Wang B, Zhang H, et al. Clinical characteristics of acute respiratory syndrome with SARS-CoV-2 infection in children in South China [published online ahead of print, 2020 Jun 24]. Pediatr Pulmonol. 2020;10.1002/ppul.24921. doi:10.1002/ppul.24921

[49] Zhong Z, Xie X, Huang W, et al. Chest CT findings and clinical features of coronavirus disease 2019 in children. Journal of Central South University(Medical sciences). 2020;45(3):236–242. doi:10.11817/j.issn.1672-7347.2020.200206 (in chinese)

[50] Zhu L, Wang J, Huang R, et al. Clinical characteristics of a case series of children with coronavirus disease 2019. Pediatr Pulmonol. 2020;55(6):1430–1432. doi:10.1002/ppul.24767

[51] Huang XP, Xie SH, Jiang LL, et al. Analysis of clinical characteristics and imaging characteristics of COVID-19 in children in Gannan region. Journal of Gannan Medical University.2020,40(03):239–242. doi:10.3969/j.issn.1001-5779.2020.03.005 (in chinese)

[52] Wu HP, Li BF, Chen X, et al. Clinical features of coronavirus disease 2019 in children aged <18 years in Jiangxi,China: an analysis of 23 cases. Chinese Journal of Contemporary Pediatrics.2020,22(05):419–424. doi: 10.7499/j.issn.1008-8830.2003202 (in chinese)

[53] Li Q, Peng X, Sun Z, et al. Clinical and imaging characteristics of children with corona virus disease 2019 (COVID-19). Radiol Practice, 2020, 35: 277–80. doi:10.13609/j.cnki.1000-0313.2020.03.007

[54] Liu J, Luo W, Deng Z, et al. Clinical and epidemiological characteristics of 91 children conformed with COVID-19. Chin J Nosocomiol, 2020, 30: 1645–9. doi:10.11816/cn.ni.2020-200550

[55] Wang KJ, Xu L,Yin Hui, et al. Clinical and CT Imaging Findings of Novel Coronavirus Pneumonia in Children. Journal of Hubei University of Medicine.2020,39(02):134–138. doi:10.13819/j.issn.2096-708X.2020.02.007 (in chinese)

[56] Yang L, Li Z, Xu H, et al. Epidemiological and clinical characteristics of 10 children with coronavirus disease (COVID-19) in Jinan City. Journal of Shandong University(Health Sciences).2020,58(04):36–39. doi:10.6040/j.issn.1671-7554.0.2020.305 (in chinese)

[57] Yin H, Wu HY, Chen J, et al. Clinical characteristics and bedside lung ultrasound findings of family-clustered pediatric COVID-19. Guangxi Medical Journal.2020,42(10):1281–1284. doi:10.11675/j.issn.0253-4304.2020.10.22 (in chinese)

[58] Zhang XG, Ma Y, Xiao JA, et al. Clinical characteristics of novel coronavirus pneumonia in children in Jinan. Journal of Shandong University(Health Sciences).2020,58(03):62–64. doi:10.6040/j.issn.1671-7554.0.2020.180 (in chinese)

[59] Fu L, Wang B, Yuan T, et al. Clinical characteristics of coronavirus disease 2019 (COVID-19) in China: A systematic review and meta-analysis. J Infect. 2020;80(6):656–665. doi:10.1016/j.jinf.2020.03.041

[60] Li LQ, Huang T, Wang YQ, et al. COVID-19 patients’ clinical characteristics, discharge rate, and fatality rate of meta-analysis. J Med Virol. 2020;92(6):577–583. doi:10.1002/jmv.25757

[61] Huang C, Wang Y, Li X, et al. Clinical features of patients infected with 2019 novel coronavirus in Wuhan, China. Lancet. 2020. 395(10223): 497–506.

[62] Chen N, Zhou M, Dong X, et al. Epidemiological and clinical characteristics of 99 cases of 2019 novel coronavirus pneumonia in Wuhan, China: a descriptive study.Lancet. 2020. 395(10223): 507–513.

[63] Li Q, Guan X, Wu P, et al. Early Transmission Dynamics in Wuhan, China, of Novel Coronavirus-Infected Pneumonia. N Engl J Med. 2020.

[64] Chen L, Liu H G, Liu W, et al. Analysis of clinical features of 29 patients with 2019 novel coronavirus pneumonia[J]. Zhonghua jie he he hu xi za zhi= Zhonghua jiehe he huxi zazhi= Chinese journal of tuberculosis and respiratory diseases, 2020, 43: E005–E005.

[65] Chen G., Wu D., Guo W., et al. Clinical and immunologic features in severe and moderate forms of coronavirus disease 2019. 2020: medRxiv 2020.02.16.20023903.

[66] Li Y, Xia L. Coronavirus Disease 2019 (COVID-19): Role of Chest CT in Diagnosis and Management. AJR Am J Roentgenol. 2020;214(6):1280–1286. doi:10.2214/AJR.20.22954

[67] Zhou S, Wang Y, Zhu T, Xia L. CT Features of Coronavirus Disease 2019 (COVID-19) Pneumonia in 62 Patients in Wuhan, China. AJR Am J Roentgenol. 2020;214(6):1287–1294. doi:10.2214/AJR.20.22975

[68] Chang TH, Wu JL, Chang LY. Clinical characteristics and diagnostic challenges of pediatric COVID-19: A systematic review and meta-analysis. J Formos Med Assoc. 2020;119(5):982–989. doi:10.1016/j.jfma.2020.04.007

[69] Liu H, Liu F, Li J, Zhang T, Wang D, Lan W. Clinical and CT imaging features of the COVID-19 pneumonia: Focus on pregnant women and children. J Infect. 2020;80(5):e7–e13. doi:10.1016/j.jinf.2020.03.007

